# X-Chromosome Association Study in Latin American Cohorts Identifies New Loci in Parkinson Disease

**DOI:** 10.1101/2023.01.31.23285199

**Authors:** Thiago P Leal, Jennifer N French-Kwawu, Mateus H Gouveia, Victor Borda, Miguel Inca-Martinez, Emily A Mason, Andrea RVR Horimoto, Douglas P Loesch, Elif I Sarihan, Mario R Cornejo-Olivas, Luis E Torres, Pilar E Mazzetti-Soler, Carlos Cosentino, Elison H Sarapura-Castro, Andrea Rivera-Valdivia, Angel C Medina, Elena M Dieguez, Víctor E Raggio, Andrés Lescano, Vitor Tumas, Vanderci Borges, Henrique B Ferraz, Carlos R Rieder, Artur Schumacher-Schuh, Bruno L Santos-Lobato, Carlos Velez-Pardo, Marlene Jimenez-Del-Rio, Francisco Lopera, Sonia Moreno, Pedro Chana-Cuevas, William Fernandez, Gonzalo Arboleda, Humberto Arboleda, Carlos E Arboleda Bustos, Dora Yearout, Maria F Lima-Costa, Eduardo Tarazona, Cyrus Zabetian, Timothy A Thornton, Timothy D O’Connor, Ignacio F Mata, the Latin American Research Consortium on the Genetics of Parkinson’s Disease (LARGE-PD)

## Abstract

Sex differences in Parkinson Disease (PD) risk are well-known. However, it is still unclear the role of sex chromosomes in the development and progression of PD. We performed the first X-chromosome Wide Association Study (XWAS) for PD risk in Latin American individuals. We used data from three admixed cohorts: (i) Latin American Research consortium on the GEnetics of Parkinson’s Disease (n=1,504) as discover cohort and (ii) Latino cohort from International Parkinson Disease Genomics Consortium (n = 155) and (iii) Bambui Aging cohort (n= 1,442) as replication cohorts. After developing a X-chromosome framework specifically designed for admixed populations, we identified eight linkage disequilibrium regions associated with PD. We fully replicated one of these regions (top variant rs525496; discovery OR [95%CI]: 0.60 [0.478 - 0.77], p = 3.13 × *10^-5^* ; replication OR: 0.60 [0.37-0.98], p = 0.04). rs525496 is an expression quantitative trait loci for several genes expressed in brain tissues, including *RAB9B, H2BFM, TSMB15B* and *GLRA4*. We also replicated a previous XWAS finding (rs28602900), showing that this variant is associated with PD in non-European populations. Our results reinforce the importance of including X-chromosome and diverse populations in genetic studies.

## 1. Introduction

Parkinson disease (PD) is the most common movement disorder and the second most common neurodegenerative disease [1,2]. PD has an estimated prevalence of 1 to 2 per 1,000 individuals and affects about 1% of the population older than 60 years old [2]. Although no singular definite answer lies behind the etiology, there is substantial evidence that PD develops from multiple factors, including genetics, age, sex, and environmental factors. The multifactorial nature of PD leads to variable clinical manifestations and response to treatment among individuals.

Sex is an important factor, not only in the development but also in the progression of PD. Studies have shown that PD affects males more frequently than females, although a recent meta-analysis showed that this difference in prevalence can be lower than expected and is population-specific [3]. Biological aging differs distinctly between males and females due to hormonal and immunological changes that females specifically experience [4–6], and this may contribute to the stark differences observed in PD risk, presentation, severity, and treatment success [7]. Even though estrogen is thought to have a protective role in PD risk [8–10], this remains unclear, as multiple studies contradict this hypothesis [11–14]. Currently, there is a huge gap in information about the cause of these differences in incidence and manifestation, including the impact of factors related to women’s health (pregnancy, menses, etc.) on the development and progression of PD [15].

Aside from sex, genetics plays a large role in PD. Genome-wide association studies (GWAS) have identified close to 100 low-penetrance variants associated with PD [16,17]. However, much of what we know about the genetic architecture of PD is based on European and Asian populations, leading to a limited understanding in other populations such as Latin Americans [18], which are the product of intensive admixture during the last 500 years between Africans, Europeans and Native Americans. The Latin American Research Consortium on the GEnetics of PD (LARGE-PD), an ongoing effort of more than 40 institutions in 14 countries across the Americas and the Caribbean, was formed to address this gap. We recently performed the first PD GWAS [19] and polygenic risk score analyses [20] in our highly admixed Latin American cohort.

Although the GWAS approach has been instrumental in the field, the genetic associations found for PD risk have been derived from autosomal variants. The exclusion of the sex chromosomes in GWAS, especially the X-chromosome (X-chr), is very common due to the challenges (e.g., poor array coverage, complex statistical analyses, etc.) that are not present for autosomal chromosomes. However the X-chr has about 155 Mb of DNA and includes about 5% of the entire genome[21,22].

The differences in prevalence between sexes, combined with studies that did not find significant genetic differences in autosomal chromosomes between PD cases in males and females [23], led to the hypothesis that genetic variants on the X-chr could explain these differences between males and females. To test this hypothesis, Le Guen et al. performed the first X-chromosome wide association study (XWAS) in PD [24]. This study identified two significant loci, one of which (rs28602900) was replicated in an independent cohort when analyzing males and females together, but found no sex-specific variants. Although this study paved the way for a better understanding of the role that X-chr may play in PD etiology, it included only individuals of European ancestry, leaving our understanding uncertain of the role of the X-chr in other populations.

Here, we show the results of the first-ever XWAS for PD in a highly diverse and admixed cohort, composed of 1,498 individuals from five countries in Latin America as part of LARGE-PD, and used an independent cohort of 1,577 admixed Latin American individuals to replicate our results.

## 2. Methods

### 2.1 Samples

#### 2.1.1 Discovery cohort: LARGE-PD dataset

For the discovery cohort, we included 1,498 individuals from Brazil, Chile, Colombia, Peru, and Uruguay recruited as part of LARGE-PD. This cohort has a mean age of 59.3±13.9 years, with 55.7% being females. After excluding all samples with missing data (n=17), our dataset was composed of 798 cases (374 females) and 683 controls (453 females). A detailed description of LARGE-PD and genotyping can be seen in Loesch et al. 2021 [19].

We carried out the analysis using all individuals together (LARGE-ALL, n=1,481) as well as by each LARGE-PD sub-cohort: Brazil (n=223), Chile and Colombia (n=359), Peru (n=710), and Uruguay (n=189). We merged Chile and Colombia due to Chile having only cases and Colombia being a population with a similar ancestry background based on PCA and ADMIXTURE inferences[19,25].

#### 2.1.2 Replication cohort: IPDGC and the Bambuí Cohort of Aging

For our replication cohort, we combined two independent cohorts, one from the International Parkinson Disease Genomics Consortium (IPDGC) and the other the Bambuí Aging Cohort Study [26]. The IPDGC Latino cohort included 155 samples,117 cases (44 females) and 38 controls (22 females) (age unavailable), ethnically-matched with LARGE-PD and genotyped with the Illumina NeuroChip Array [27].

This IPDGC cohort was combined with data from the Bambuí to increase the sample size. This dataset is composed of 1,422 individuals, representing 82% of the residents in the city of Bambuí (Brazil) older than 60 years old at the baseline year (1997) [28]. A detailed description of Bambui cohort and genotyping can be seen in Kehdy et al [29].

To include the Bambuí dataset, we calculated the autosomal principal component analysis (PCA) for the IPDGC + Bambuí with European and African populations from 1000 Genomes (YRI and IBS respectively [30]) and Native American (dataset from the Tarazona Lab [29]) as parental references. After this, we calculated the mean and standard deviation (sd) for IPDGC samples for the first 10 PCs and removed all Bambui individuals that were _+_2 sd from the IPDGC mean in any PC. The Bambuí cohort contains mostly healthy controls but also a few idiopathic PD [31]. In our study we included 10 idiopathic PD as cases. Individuals without PD diagnosis were added as controls (275 males, 509 females)

Together, our replication cohort included 949 individuals, of which 127 are cases (48 females) and 822 controls (531 females). Details about the IPDGC + Bambuí dataset can be found in the Supplementary information (SI) (**SI section:** Bambuí sample selection)

### 2.2 Quality control, admixture Analysis, and imputation

#### 2.1.1 Basic quality control

We first performed a basic quality control (QC) procedure (**Figure S1**) using PLINK v1.90 [32] in which we removed unaligned, 100% heterozygous, and ambiguous variants for the discovery and replication cohorts. Next, we excluded variants and individuals with more than 10% missing data, splitted the pseudo-autosomal data from X-chr, and removed individuals whose genetic sex, inferred by PLINK, differed from the self-reported sex. Furthermore, we changed the reference genome from hg37 to hg38 using an in-house script.

#### 2.2.2 Le Guen et al. 2021 QC process

As reference, we utilized the QC and harmonization pipeline that Le Guen and collaborators developed, but we implemented some modifications as their pipeline was specific for a homogeneous ancestry cohort [24], and LARGE-PD consist of highly admixed individuals.

To apply the Le Guen et al. pipeline to a non-homogeneous population, and mitigate the risk of false positives caused by population structure, our pipeline was composed of three main steps: (i) Autosomal QC **(Figure S2)**, (ii) X-chromosome QC **(Figure S3)**, and (iii) Population structure analysis **(Figure S4)**. The differences will be explained in each step.

##### 2.2.2.1 Autosomal QC

First, we removed individuals with missing covariates and variants that are: (i) located in structural variants using TriTyper [33], (ii) duplicated, (iii) monomorphic, (iv) potential probe sites (defined as variants in which the probe may have variable affinity due to the presence of other SNPs within 20 bp and with MAF above 1%), or (v) have failed the Hardy-Weinberg Equilibrium (HWE) exact test (*p* < 10^−5^ in controls). Next, we removed individuals with > 10% and variants with more than 5% of missing genetic data and inferred the relatedness between included individuals using KING [34]; those at greater than a second degree level (kinship coefficient > 0.0884) were removed using NAToRA [35] (**Figure S2**). Unlike the Le Guen et al. pipeline, samples were not removed based on ancestry.

##### 2.2.2.2 X-Chromosome QC

We removed any individual excluded in the Autosomal QC step and performed the same steps from the Autosomal QC on X-chr variants, except for the relatedness calculation and removal. Specific to the X-chr, we removed variants if they (i) were heterozygous variants in males, (ii) had differential missingness between cases and controls (p-value <10^−5^), (iii) failed the HWE exact test (p-value <10^−5^) in the female controls, (iv) had differential missingness and MAF between males and females (p-value <10^−5^). Next, we set males as hemizygous and phased our data using 1000 Genomes Project [30] as reference and the flags “--allowRefAltSwap -- keepMissingPloidyX” in Eagle (v2.4.1)[36](**Figure S3**).

We performed all QC steps described for Brazil, Chile+Colombia, Peru, Uruguay, LARGE-ALL, and IPDGC + Bambui.

### 2.3 Imputation

Imputation for all cohorts was done with the TOPMed Imputation Server [37] and a local version of the TOPMed Imputation Panel using a subset of Freeze 10b [38]. After a pilot test, we opted to use the local version because the results were slightly better than those obtained with the Imputation Server. We also compared the imputation quality between autosomal and X-chr and observed a decrease in imputation quality for the X-chr compared to the autosomal (**Figure S5, Table S1**). We chose to use only variants with a high imputation quality score (r^2^ > 0.8). More details about the imputation tests can be found in SI (**SI: section Imputation Tests**).

### 2.4 Population structure control

For population structure control, we used X-chr principal component analysis (X-PCA) instead of the autosomal PCA due to the low correlation between them (**SI: section X-chromosome principal component analysis**).

We performed 10 X-PCAs using the QCed genotyped data using the GENESIS package [39] and we removed all outlier samples, defined as ± 3 sd in any PC for males and females. We then merged non-outlier males and females to generate the Both dataset (males and females combined). With this new dataset, we performed the X-PCA and removed outliers from the Both dataset (**Figure S4, Figure S6**). The number of samples and variants per dataset are presented in **Table 1**.

**Table 1:** Information about the different databases used in this study. We calculated the number of independent tests and the p-value threshold (PVT) for each dataset. We use the smallest value as the LARGE-Meta cutoff.

|  |  | Discover Phase |  |  |  |  | Replication Phase |
| --- | --- | --- | --- | --- | --- | --- | --- |
|  |  | Brazil | Chile and Colombia | Peru | Uruguay | LARGE-ALL | IPDGC + Bambuí |
| # of variants | Genotyped | 15,831 | 16,109 | 14,844 | 14,388 | 17,860 | 3,599 |
|  | Genotyped and imputed | 562,354 | 634,449 | 438,816 | 474,046 | 763,375 | 373,270 |
| Males | # samples | 102 | 152 | 288 | 62 | 576 | 260 |
|  | # cases | 87 | 68 | 214 | 23 | 375 | 72 |
|  | PVT genotyped | 3.73E-04 | 3.10E-04 | 1.47E-04 | 3.56E-05 | 1.00E-04 | N/A |
|  | PVT imputed | 3.22E-04 | 2.77E-04 | 1.15E-04 | 2.33E-05 | 6.65E-05 | N/A |
| Females | # samples | 95 | 172 | 364 | 108 | 754 | 345 |
|  | # cases | 47 | 79 | 184 | 24 | 340 | 34 |
|  | PVT genotyped | 4.70E-04 | 3.18E-04 | 2.22E-04 | 4.20E-05 | 1.46E-04 | N/A |
|  | PVT imputed | 4.26E-04 | 2.99E-04 | 2.31E-04 | 3.99E-05 | 1.33E-04 | N/A |
| Both | # samples | 175 | 295 | 602 | 149 | 1,193 | 544 |
|  | # females | 73 | 144 | 314 | 87 | 618 | 284 |
|  | # cases | 117 | 138 | 370 | 42 | 658 | 100 |
|  | PVT genotyped | 3.31E-04 | 2.37E-04 | 1.58E-04 | 3.28E-05 | 1.10E-04 | N/A |
|  | PVT imputed | 3.30E-04 | 2.41E-04 | 1.77E-04 | 3.08E-05 | 8.43E-05 | N/A |
| PVT:p-value threshold |  |  |  |  |  |  |  |

### 2.5 Association study

Differences in allelic frequencies caused by historical/demographic variation could increase our false-negative or false-positive results. Due to our small sample size, to avoid a significant loss in statistical power for the association analyses, we used two approaches. To ensure the largest sample size possible for analyses, we used the LARGE-ALL dataset, whereas to control for the risk of heterogeneity caused by historical/demographic events, we used a meta-analysis approach, called here as LARGE-Meta.

For LARGE-ALL, we merged the five sub-cohorts and performed all the QC steps, imputation, population structure control, and regression in a single run. For LARGE-Meta, we first performed the QC steps, imputation, population structure control, and regression by sub-cohort followed by a meta-analysis using the GWAMA software [40] (**Figure S7**).

For all datasets, we performed regression for three datasets: (i) Male, (ii) Female, and (iii) Both (regression with males and females merged). We also performed a meta-analysis using sex-differentiated and sex heterogeneity implemented on GWAMA [40], called Male+Female by Le Guen and collaborators (**Figure S6**).

All regression analyses were performed using Firth’s logistic regression in PLINK2 [41]. In each of our models, we used age (when available), sex, and X-PCs as covariates. Sex-stratified analysis involved PC1 to PC10, while for the Both dataset we used PC2 to PC10 because, in this dataset, PC1 separates males and females. We used the PLINK2 default X-chromosome coding, which males and females are both on a 0 to 2 scale.

#### 2.6 X-Chromosome p-value threshold

We calculated the X-chr p-value threshold (PVT) **(Table 1)** based on the number of independent tests for each dataset (**SI: section X-Chromosome significance threshold**). We did not used the GWAS PVT (p < *5□10^−8^*) because the Genome-Wide analysis has more independent tests than X-chr analysis.

#### 2.7 Gene expression studies

We took the top variants from replicated regions and used GTEx Portal [42] to identify the genes for which these variants act as eQTL. We built a list with all eQTLs searching the rs ID in the portal (https://gtexportal.org/home/) and all genes present in “Single-Tissue eQTLs” section were included in our list of genes.

After this, we accessed through AMP-PD (https://amp-pd.org/transcriptomics-data) the whole-blood time progression gene expression data (gencode v29) from PPMI (https://amp-pd.org/unified-cohorts/ppmi) and extracted the RNA counts for our list of genes from baseline point (0 month time point).

To investigate if our variants are associated with differences in gene expression, we use a t-test to assess whether there is a statistical difference between the RNA counts of cases and controls segregating by sex and self-reported ethnicity (Hispanic or Latino and non-Hispanic or Latino).

## 3. Results

### 3.1 Association results

The association study revealed that 86 candidate variants were significantly associated based on the X-chr PVT (**Figure 1, Figure S8, Table 1, Table S2-S5)**.

**Figure 1:**
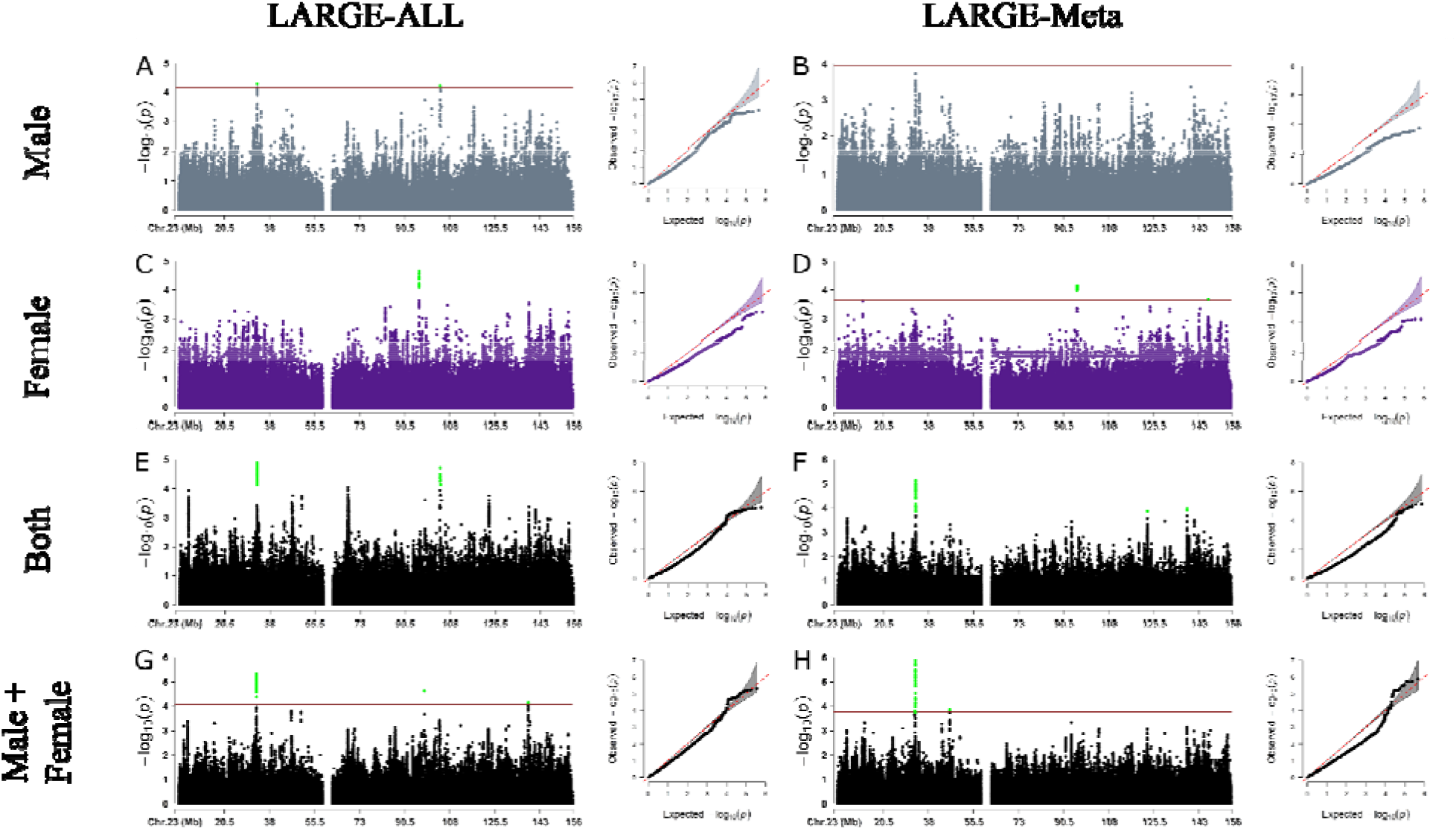
Results from X-chromosome Wide Association Study in LARGE-PD dataset for the discovery phase using LARGE-ALL (left) and LARGE-Meta (right). Each panel shows the Manhattan and Q-Q Plot for Males (A and B), Females (C and D), Both (E and F), and Male + Female (G and H). The significance threshold was calculated based on the number of independent tests. Because we were not able to calculate the cutoff value for the LARGE-Meta, we opted to use the same cutoff value for the Both dataset.

We observed that the Females analysis was the most underpowered based on QQ plots (**Figure 1C and D**), even though it has more samples than the Males analysis. We hypothesized that this could be caused by the fact that female heterozygous variants could act as noise because we cannot infer what X-chr was inactivated in each cell. To test this hypothesis, we treated the heterozygous variants as missing data and performed the regression. In general, the inflation rate was better, but this resulted in the loss of ∼ 60% of variants in all datasets due to an increase in the number of monomorphic variants (**Figure S9**), thus we decided against this approach.

Next, we analyzed all XWAS summary statistics in all four different datasets (Male, Female, Both, and Male+Female) using the LARGE-ALL dataset and LARGE-Meta approach. Using the LDMatrix [43] and Admixed American Populations as reference, we then found the 86 variants identified in the association study to be in eight regions with high linkage disequilibrium (LD) (**Figure S10**). All variants that reached statistical significance (**Table S2-S5**) have not been previously reported.

### 3.2 Replication using the IPDGC + Bambui cohort

Using the IPDGC + Bambuí dataset, 19 variants in two high LD regions achieved statistical significance: (i) chrX:103929276-104031236 (R^2^ > 0.617, D’ = 1) and (i) chrX:95074293-95823918 (R^2^ > 0.84, D’ = 1) (**Table S6**).

The region chrX:103929276-104031236 (**Figure S12**), represented by rs525496 (**Figure S11**), was discovered in LARGE-ALL Both (OR [95% CI]: 0.60 [0.48, 0.77], p: 3.13×10 ^5^), and it was replicated in Males (OR [95% CI]: 0.60 [0.37, 0.98], p: 0.0421). Besides that, the p-values for Both dataset on the replication cohort were near the PVT (p ranging between 0.062-0.088). The region includes three genes, *TMSB15B, H2BW1*, and *H2BW2*, and it is close to *SLC25A53, FAM199X*, and *ZCCHC18* genes.

The region chrX:95074293-95823918, represented by rs112103361 (**Figure S13**), is an intergenic region and it was discovered in Male analysis in LARGE-ALL (OR [95%CI]: 1.65 [1.31, 2.08], p: 2.24×10 ^5^) and LARGE-Meta (1.66 [1.29, 2.12], p: 6.8×10 ^5^) and it achieved statistical significance in the replication cohort in Males (OR [95%CI]: 0.53 [0.32, 0.89], p: 0.017) and in Male+Female analysis (0.59 [0.39, 0.89], p= 0.012). However, the effect directions for the replication were opposite to those in the discovery results. We were also able to replicate one of the variants from Le Guen et al., rs28602900, with the same direction (OR > 1) (**Figure S14**).

This variant is located in a high gene density region that includes *RPL10, ATP6A1, FAM50A* and *PLXNA3* and also was replicated in the original paper. The other variant, rs7066890, was not present in our dataset due to low imputation score (r^2^ < 0.8).

### 3.3 Differences in RNA expression on eQTL genes

Using the GETx Portal [42] we observed that rs525496 (and the variants in the validated region) acts as eQTL for several genes in different tissues (**Table S7**). We have four genes in brain tissues: (i) *TMSB15B*, (ii) *LL0XNC01-116E7*.*1* and (iii) *GLRA4*, present in cerebellum, and (iv) H2BFM in brain - nucleus accumbens (basal ganglia). Other genes in non-brain tissue include *H2BFM, FAM199X, RAB9B,ZCCHC18, LL0XNC01-116E7*.*2*, and *LL0XNC01-240C2*.*1*.

We tried to download the PPMI data for all mentioned genes, but only information for*GLRA4, RAB9B, H2BFM, ZCCHC18*, and *FAM199X* was available. After the t-test, we observed statistical differences for *GLRA4* in females in both ethnic groups and for *RAB9B* and *H2BFM* in non-Hispanic/Latinos in Females and Males (**Figure S15**). However PPMI has only data for 130 Hispanic/Latinos compared to 1,139 non-Hispanic/Latino.

## 4. Discussion

GWAS frequently exclude analysis of the X-chromosome, resulting in a lack of understanding about the role in disease of ∼ 5% of the human genome. In addition, population diversity has been inadequately represented in genetic studies, with most studies including only individuals of European ancestry [44,45]. Both are true in PD, where only one XWAS has been performed, including only individuals of European ancestry [24]. Here, we performed the first XWAS for PD in a Latin American cohort and shared a novel pipeline for XWAS analysis in an ancestrally diverse population. We identified 86 variants with statistical significance across eight regions with high LD, and two regions with high LD achieved statistical significance in an independent cohort.

The first region, chrX:103929276-104031236, was discovered in the Both dataset and it was replicated in Males. This region has several genes: *TMSB15B* (thymosin beta 15B), *H2BW1* (H2B histone family member W, Testis Specific), and *H2BW2* (H2B histone family member M), and it is near *SLC25A53* (solute carrier family 25 Member 53), *ZCCHC18* (zinc finger CCHC-type containing 18) and *FAM199X* (family with sequence similarity 199, X-linked). Moreover, analysis of the GTEx project revealed that the top hit (rs525496) acts as eQTL for several genes in different tissues **(Table S7)**. We found eQTLs for 4 different genes in brain: (i) *TMSB15B*, (ii) *LL0XNC01-116E7*.*1* and (iii) *GLRA4*, in cerebellum, and (iv) *H2BFM* in brain - nucleus accumbens (basal ganglia). None of these four genes have been previously associated with PD. *GLRA4* is a glycine receptor with some studies showing that glycine may have neuroprotective properties [46,47]. Furthermore, a deletion of the chromosomal region including *GLRA4* was associated with craniofacial anomalies, intellectual disability, and behavioral problems [48]. This variant is also an eQTL for *RAB9B* gene in cells (cultured fibroblasts), a non-brain tissue.

Several studies have shown a relationship between the Ras analog in brain genes (*RAB*) in the pathogenesis of PD [49,50]. Although none of these studies directly mention *RAB9B*, they did investigate the role of *RAB9A*, a gene paralog of *RAB9B*. We also investigated eQTL gene expression and observed statistical differences in the RNA expression of the genes *GLRA4, RAB9B*, and *H2BFM* between cases and controls segregated by sex and ethnicity in the PPMI dataset.

The second region, chrX:95074293-95823918, was discovered in the Males analysis, and it was statistically significant in Males and in Male+Female replication. This region is inside an intergenic region. Furthermore, none of the variants in this region are *cis* or *trans* eQTLs according to the GTEx Portal [42]. In addition, the direction of effect is opposite between the discovery (OR > 1) and replication (OR < 1), suggesting this could be a spurious result.

However, this could also be caused by differences in effects between populations. We observed a difference in ancestry between our replication cohort and our discovery cohort. The allele frequency for the top variant in this region (rs112103361) in IPDGC + Bambuí shows it is closer to Brazil (most European LARGE-PD sub-cohort) and European (non-Finnish) cohort from gnomAD than to other populations from LARGE-PD or Latinos/Admixed from gnomAD (**Table S8**). Thus more studies are needed in order to understand a possible role of this region in PD.

In addition to the identification and validation of PD associated regions on the X-chr, we observed a stark difference in the quality of imputation between the X-chr and autosomes. This is important because such differences may indicate the need for adaptations in current imputation methodologies. We also observed a low correlation between autosomal and X-chr PCA, suggesting the PCA of autosomal chromosomes should not be used to control for population structure in studies investigating associations in the sex chromosomes, at least in Latin American individuals. This low correlation may be due to the fact that recombination on the X-chr only occurs in females, so evolutionary forces exert a greater effect on this chromosome.

Along with the study by Le Guen and collaborators, we have highlighted the potential role of variants in the X-chr in PD etiology. Our study also emphasized the importance of analyzing diverse ancestral backgrounds. Many genetic factors are associated with ancestry, thus studies not including individuals of diverse ancestry may result in novel loci being overlooked. However analysis of admixed populations requires complex quality and harmonization pipelines. We modified the harmonization pipeline of Le Guen and collaborators and implemented all steps to conduct an XWAS in admixed populations. All code is publicly available at https://github.com/MataLabCCF/XWAS/.

Our study has some limitations. The first is our sample size. We started our analysis with 1,498 samples, a small number of samples for an association study. This problem was aggravated when considering that some analyses were performed segregating by sex and site, which further decreased our sample size. We tried to minimize this limitation using Firth’s regression. Another limitation is the imputation performance as previously mentioned. To avoid false-positives, we chose to use only variants with high imputation quality. Another limitation is the lack of Latin American replication cohorts, especially with Native-American ancestry genetic background.

In conclusion, our work provides evidence for new loci on the X-chromosome in an admixed population using a novel pipeline specific for a heterogeneous admixed population.

Furthermore, we validated in a Latin American cohort one of the variants identified by Le Guen and collaborators. We identified new variants that act as eQTL to several genes, including GLRA4 which has been previously associated with neurodegenerative diseases, although these findings require further investigation.

## Data Availability

All data produced in the present study are available upon reasonable request to the authors

## Acknowledgments

We thank all of the individuals who participated in LARGE-PD. We also want to thank all the support staff at the different Latin American sites for their efforts and support building this incredible resource.

The authors thank Cassandra Talerico, Ph.D., a salaried employee of the Cleveland Clinic, for assistance with manuscript review and editing. The authors thank Dr. Maira Tonidandel and Dr. Paulo Caramelli for their help with PD cases in the Bambuí cohort.

## Author roles

**Conceptualization:** TPL, SCR, IFM

**Methodology:** TPL, SCR, MHG. IFM

**Software:** TPL

**Validation:** TPL, MHG

**Formal analysis:** TPL

**Investigation:** TPL, MIM, EAM, MCO, LT, CC, EHS, ACM, ED, VT, HBF, CRR, AFSS, BLSL, CVP, MJDR, FL, SMM, PCC, HA, IFM

**Resources:** JNFK, VBP, VR, AL, VB, HBF, CVP, MJDR, GA, HA, CEAB, DMY, ETS, CPZ, IFM

**Data Curation:** MHG, ES

**Writing - Original Draft:** TPL, SCR

**Writing - Review & Editing:** SCR, MHG, VBP, ARVRH, MCO, DL, ESC, HBF, CRR, AFSS, BLSL, CVP, FL, PCC, CPZ, TAT, TDO, IFM

**Visualization:** TPL

**Supervision:** MHG, TDO, IFM

**Project administration:** PM, FL, IFM

**Funding acquisition:** MJDR, CPZ, TAT, IFM

