## Supplementary material for "X-Chromosome Association Study in Latin American Cohorts Identifies New Loci in Parkinson Disease": Si text

**Bambuí sample selection**

The Bambuí Aging Cohort Study is composed of 1,422 individuals who were 82% of the residents in the city of Bambuí (Brazil) older than 60 years old at the baseline year. This dataset consists of admixed individuals whose average ancestry is 78.5% European, 14.7% African, and 6.7% Native American [[1]](https://paperpile.com/c/fnk5rp/wden). The inclusion of these data without any type of selection criteria could generate noise because it could change the genetic profile of the validation cohort, especially because the Bambuí dataset has more than 9 times the number of samples in the IPDGC dataset.

To merge IPDGC and Bambuí datasets, we performed the basic quality control procedure in both datasets (see Main article, section Basic Quality Control), selected the common variants between the IPDGC and Bambuí datasets, and merged them using PLINK 1.90 [[2]](https://paperpile.com/c/fnk5rp/4aef).

To select the Bambuí samples, we calculated the autosomal PCA for the IPDGC + Bambuí with European and African population from the 1000 Genomes Project [[3]](https://paperpile.com/c/fnk5rp/lAkl) (Iberian populations in Spain and Yoruba in Ibadan, Nigeria) and the Native American dataset from the Tarazona Lab ([[1,4]](https://paperpile.com/c/fnk5rp/92gQ+wden) as parental references. After this, we calculated the mean and sd for IPDGC samples for the first 10 Principal Components (PCs) and removed all Bambui samples that were $\pm$2 sd from the mean in any of these PCs (**Figure S16**).

The Bambuí contains identified PD cases [[5]](https://paperpile.com/c/fnk5rp/SImc) that have different causes. The most common is idiopathic PD (n=39) followed by drug-induced Parkinsonism (n=32), vascular Parkinsonism (n=13), post-traumatic Parkinsonism (n=1) and multiple system atrophy (n=1). We removed all non-idiopathic PD from the previously selected samples and included all idiopathic PD cases as cases in our replication cohort.

In total, our replication cohort has 949 individuals.

### **Imputation Tests**

We imputed all datasets (LARGE-ALL, Brazil, Chile and Colombia, Peru, Uruguay and IPDGC) using the TOPMed Imputation Server [[6]](https://paperpile.com/c/fnk5rp/kLFn) and a subset of TOPMed Freeze 10b (**Table S9**) [[7]](https://paperpile.com/c/fnk5rp/XaSn).

To determine which imputation panel better fit for our dataset, we extracted all EmpR and EmpRsq statistics provided by Minimac3 [[8]](https://paperpile.com/c/fnk5rp/QNWe). The EmpR calculates the correlation between the genotyped variant and the imputed dosages that were calculated after hiding all known genotypes of a specific variant. The EmpRsq is the square of this correlation.

After comparing both datasets, we used the TOPMed Freeze 10b panel because it had a slight improvement in EmpR and EmpRsq compared to the TOPMed Imputation server (**Table S10**, **Figure S17**), especially on the Peru dataset, our cohort with the largest number of samples.

We then compared the quality of imputation between autosomes and the X chromosome. For this, we imputed all chromosomes using the TOPMed imputation Server and extracted EmpR and EmpRsq for all chromosomes (**Figure S5** and **Table S10**). After the analysis, we concluded that the imputation process on the X chromosome is worse than on the autosomal chromosomes.

This validates our approach of using an autosomal-specific QC pipeline and an X chromosome-specific pipeline. The need for specific pipelines may be due to the fact that the X-chromosome has a different dynamic and rate of recombination, so evolutionary events have a greater impact on the X-chromosome, when compared to autosomes. Furthermore, when we looked at the analyses of the X-chromosome separated by country, we observed that the populations from Peru had the worst imputation performance. We believe this is caused by the lack of samples from populations from South America in the TOPMed imputation panel.

### **X-Chromosome principal component analysis**

To determine whether the X chromosome PCA correlated with the autosomal PCA, we performed X chromosome and autosome PCA for the LARGE-PD datasets (LARGE-ALL, Brazil, Chile + Colombia, Peru, and Uruguay). To perform the X chromosome PCA (X-PCA), we removed variants in linkage disequilibrium using PLINK2 [[9]](https://paperpile.com/c/fnk5rp/X0AWz) and the flags “--indep-pairwise 200 50 0.2”, and we calculated 10 PCs to autosomal and X chromosomes using the GENESIS package [[10]](https://paperpile.com/c/fnk5rp/vCNY) for male, female and both sexes datasets.

We calculated the correlation using male and female datasets for each cohort. We did not calculate the correlation using the both sexes datasets because the PC1 for the X chromosome separates males and females, something that does not occur for autosomal chromosomes. We believe that this occurs because in our dataset, males are hemizygous. We plotted the pairwise correlation of the first 5 PCs for male and female data and concluded that the autosomal PCA is not sufficiently correlated with the X chromosome PCA to be a set of covariates for XWAS (**Figure S18 – S27**).

### **X-Chromosome significance threshold**

To select the candidate variants we calculated an X-chromosome significance threshold instead of using the Genome-Wide significance threshold (p-value < $5x{10}^{-8}$) because this threshold aims to avoid statistically significant false-positive results that can occur by chance when multiple tests are performed at the level of 0.05, considering an experiment on the whole genome has more tests than when analyzing just the X-chromosome.

Our significance threshold was obtained based on 0.05 divided by the number of effective tests ( $N_{eff}$) [[11]](https://paperpile.com/c/fnk5rp/BYM6), [[12]](https://paperpile.com/c/fnk5rp/ze27). The $N_{eff}$ is calculated using the equation $N_{eff}= N^{2}/(\sum_{i =1}^{v} \sum_{j = 1}^{v} L_{ij})$, in which N is the number of samples on the dataset, $L$ is the $R^{2}$ correlation coefficient between all $(v)$ variants present on the datasets ($L$ is a matrix with size $v$ by $v$) and $i$ and $j$ are the matrix indexes.

We used PLINK (v1.90) to calculate the $L$ matrix between all variants (genotyped and imputed) using the flags “--r2 square gz yes-really”.

Each dataset has its own cutoff because each dataset has a different linkage disequilibrium matrix, number of variants, and sample size. All cutoffs are presented in **Table 1**.
