## Supplementary material for "X-Chromosome Association Study in Latin American Cohorts Identifies New Loci in Parkinson Disease": SI Figures

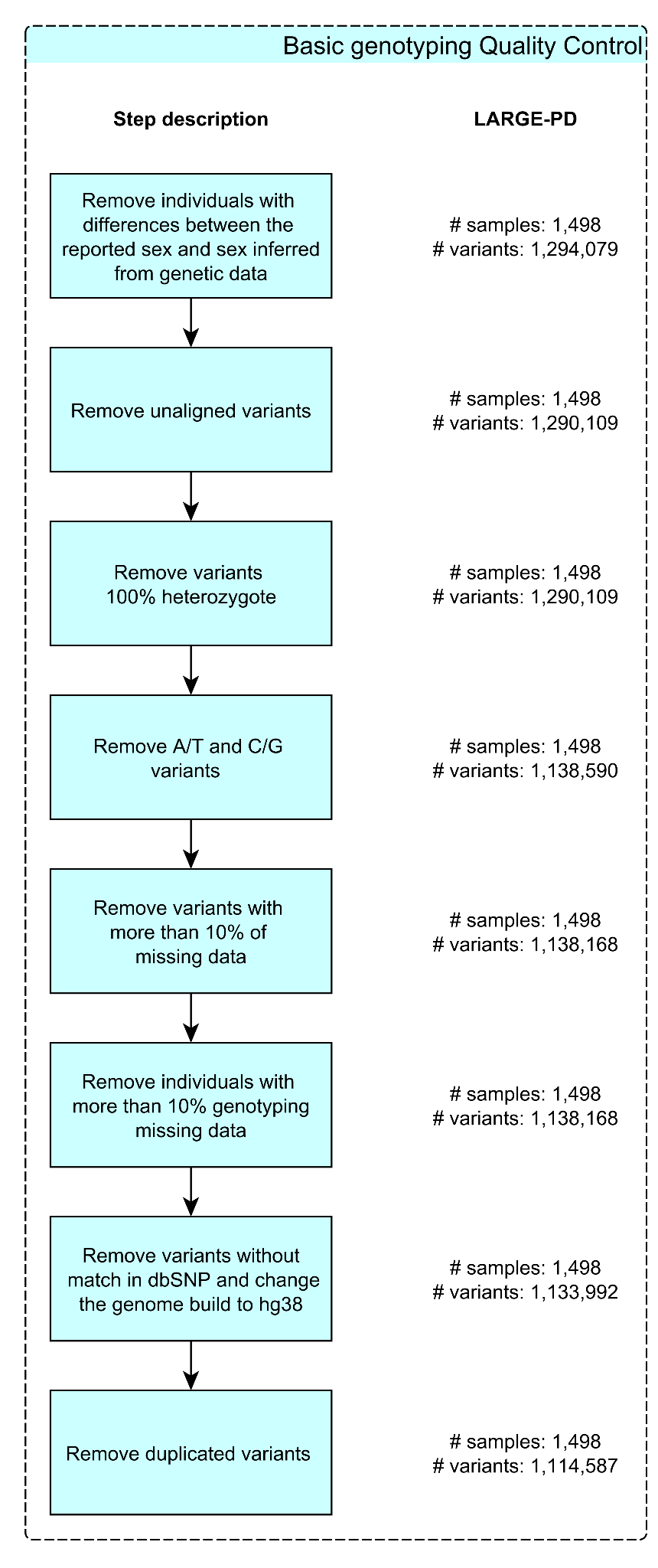

##### Figure S1: Fluxogram for basic genotyping quality control steps with the number of samples and variants per step

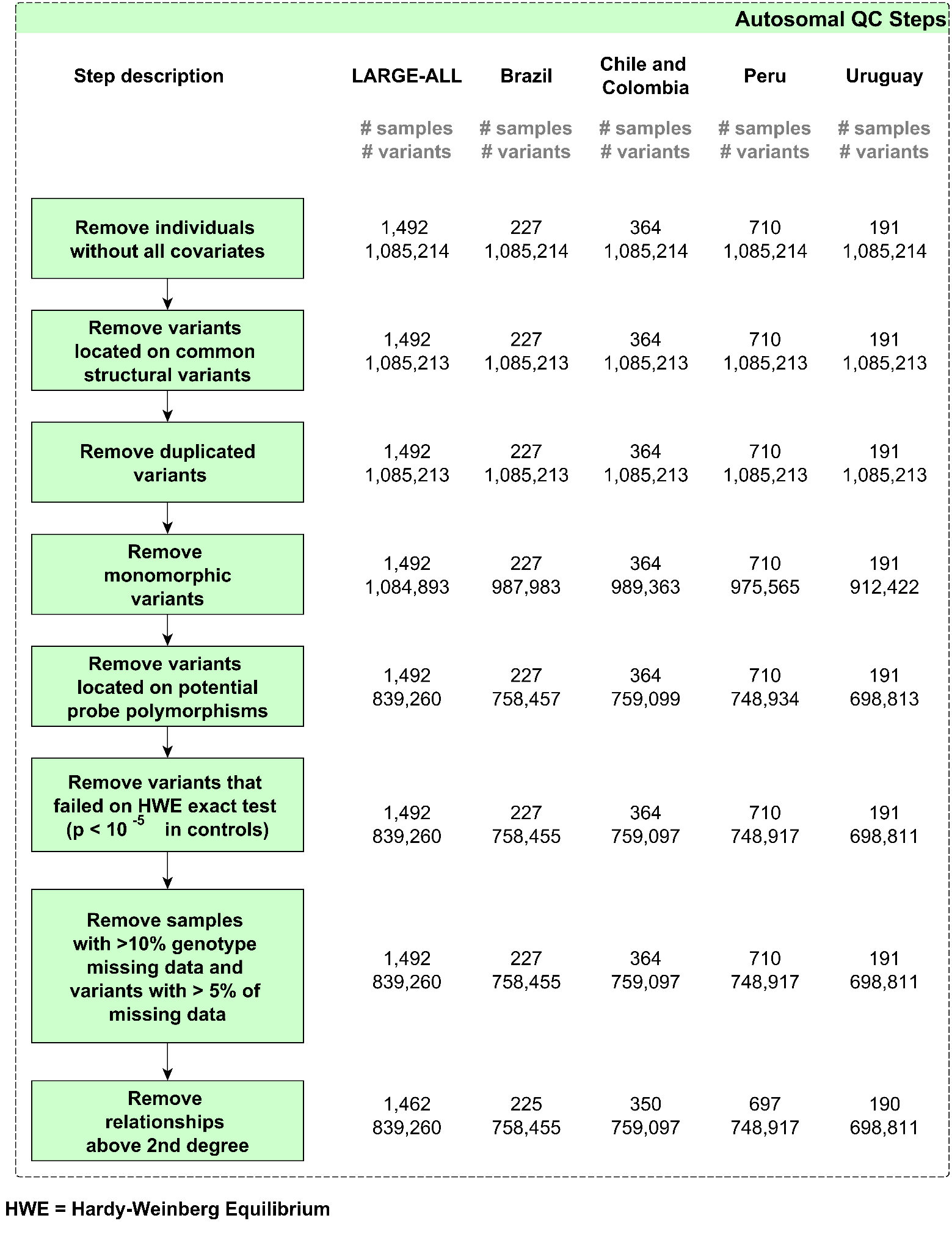

##### Figure S2: Fluxogram for autosomal QC with the number of samples and variants per step and dataset

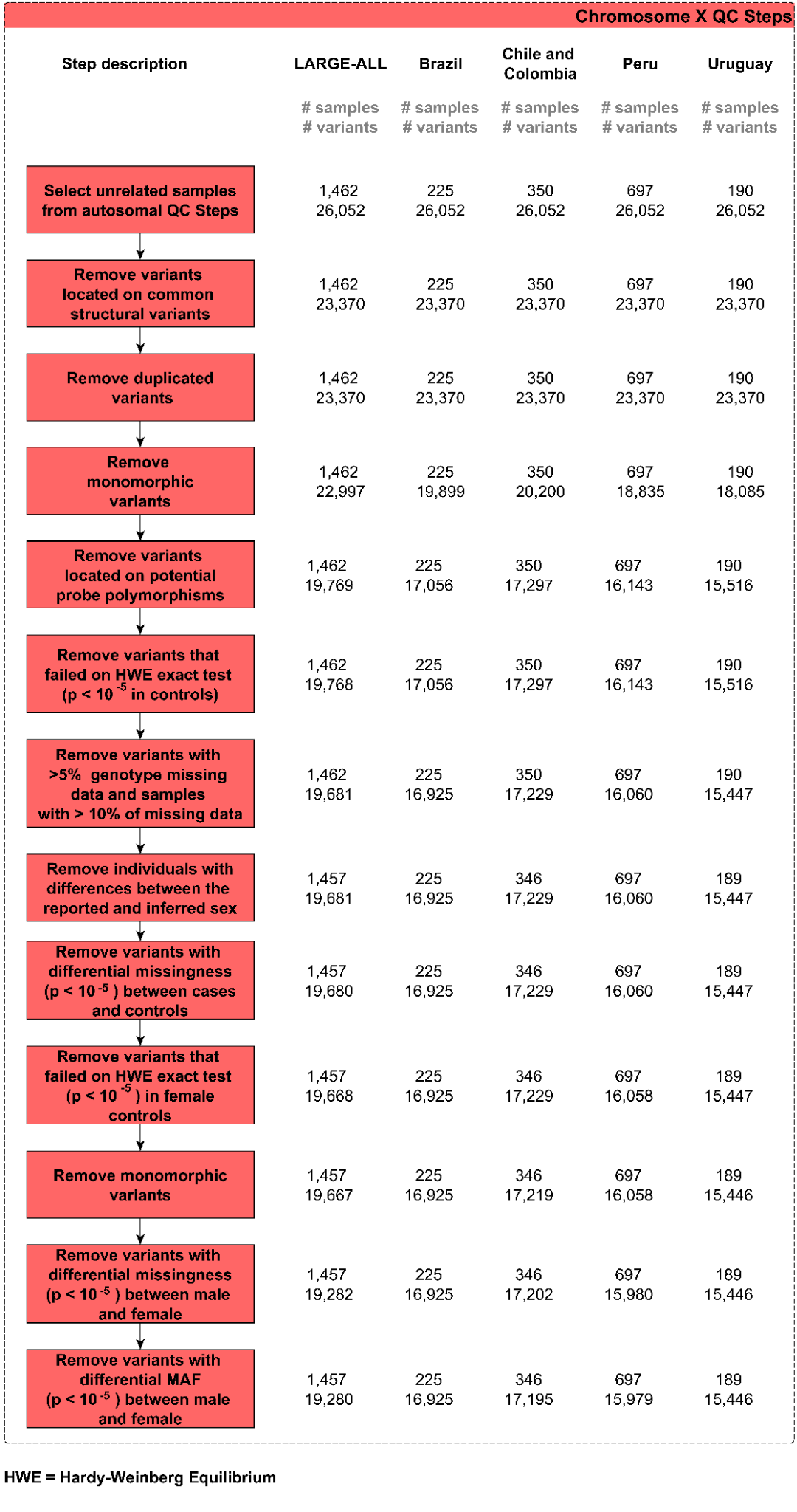

##### Figure S3: Fluxogram for chromosome X quality control with the number of samples and variants per step and dataset

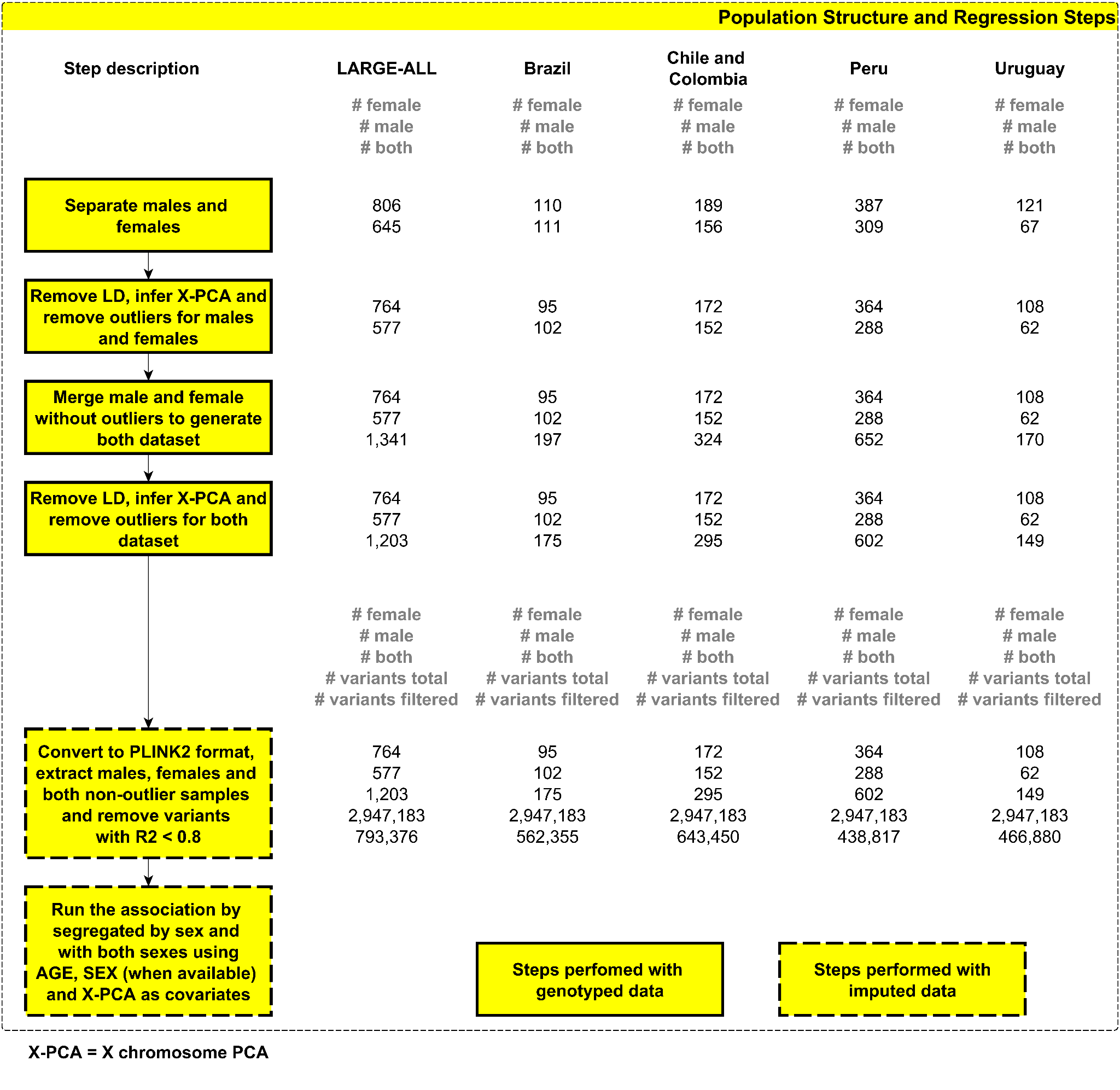

Figure S4: Fluxogram for Population Structure and regression with the number of samples and variants per step and dataset

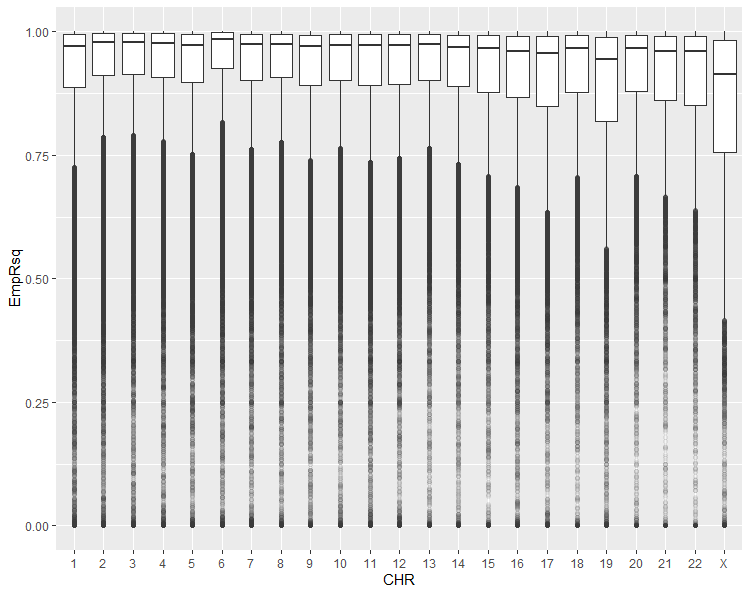

##### Figure S5: The EmpRsq per chromosome using TOPMed imputation server. We can observe that the X Chromosome has the worst EmpRsq of all chromosomes

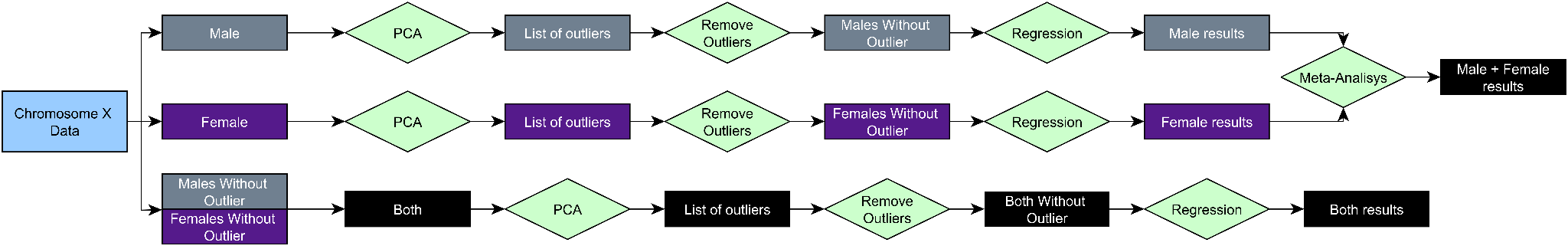

##### Figure S6: Fluxogram for Male, Female, Both and Male+Female results

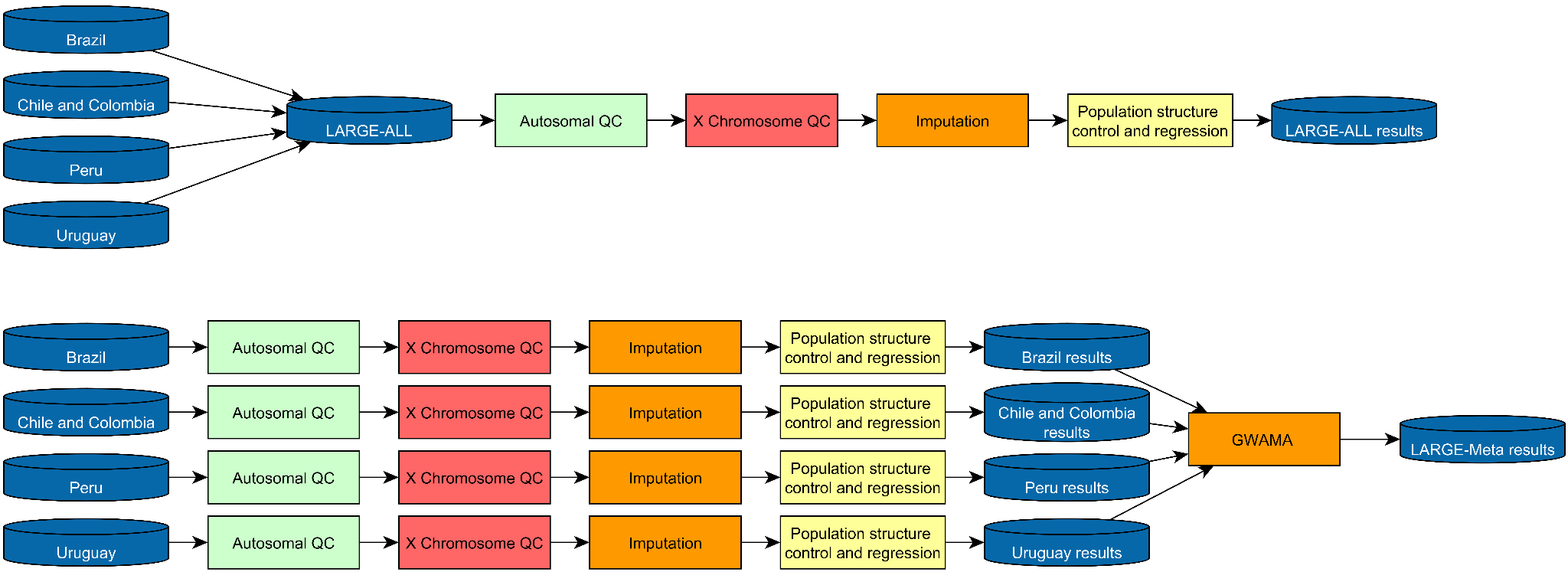

##### Figure S7: Fluxogram with LARGE-ALL and LARGE-Meta path

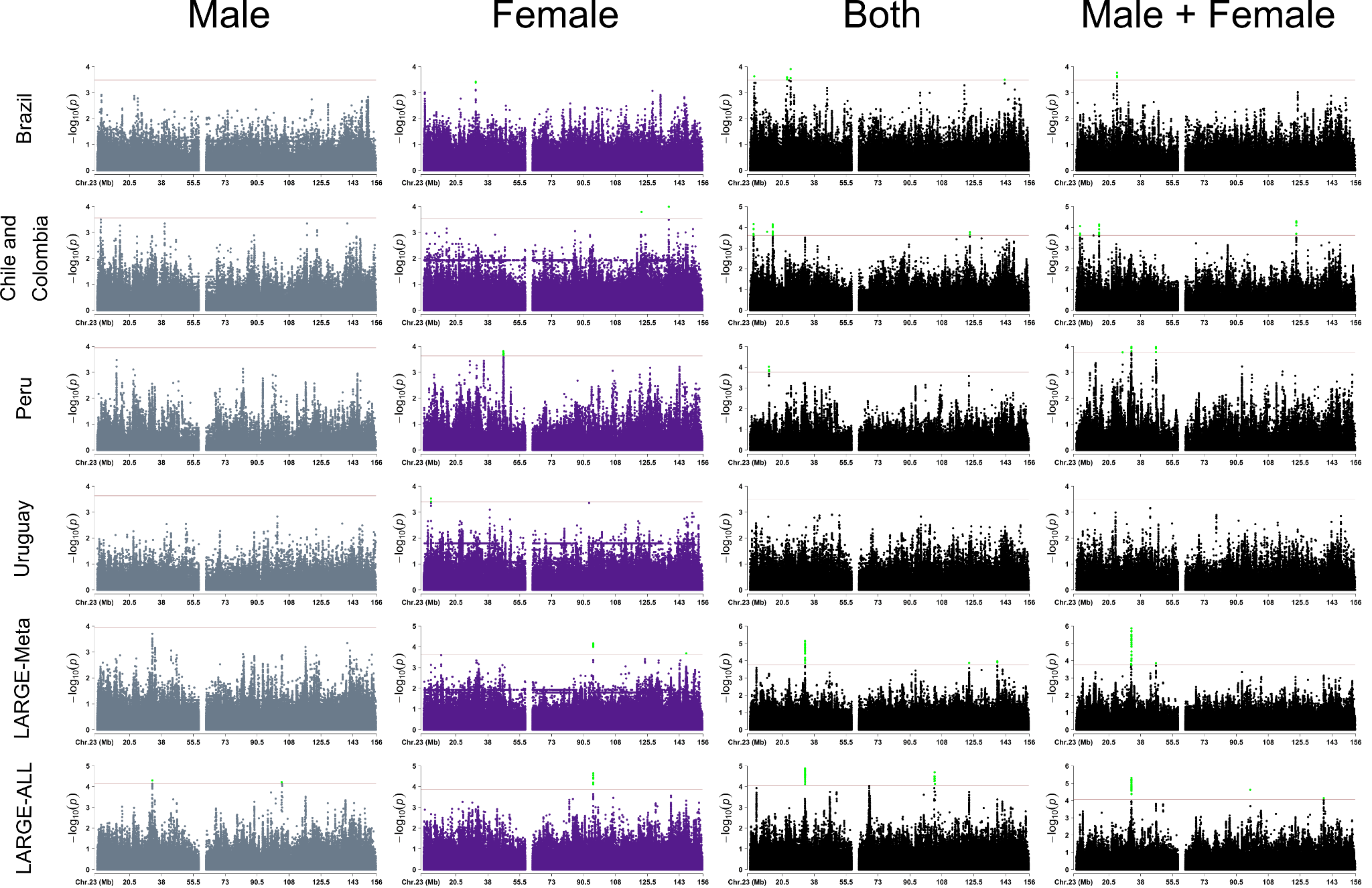

Figure S8: Manhattan plot for the regression per dataset. We present the Manhattan plots for Brazil, Chile and Colombia, Peru, Uruguay followed by the meta-analysis (LARGE-Meta) between these four datasets. The last line is the Manhattan Plot for the LARGE-ALL dataset, a dataset created by the merging of these 5 cohorts. The significance threshold is the black line on the Manhattan plots. Because we were not able to calculate the cutoff value for the meta-analysis, we opted to use the lowest cutoff value between all datasets used on the meta-analysis.

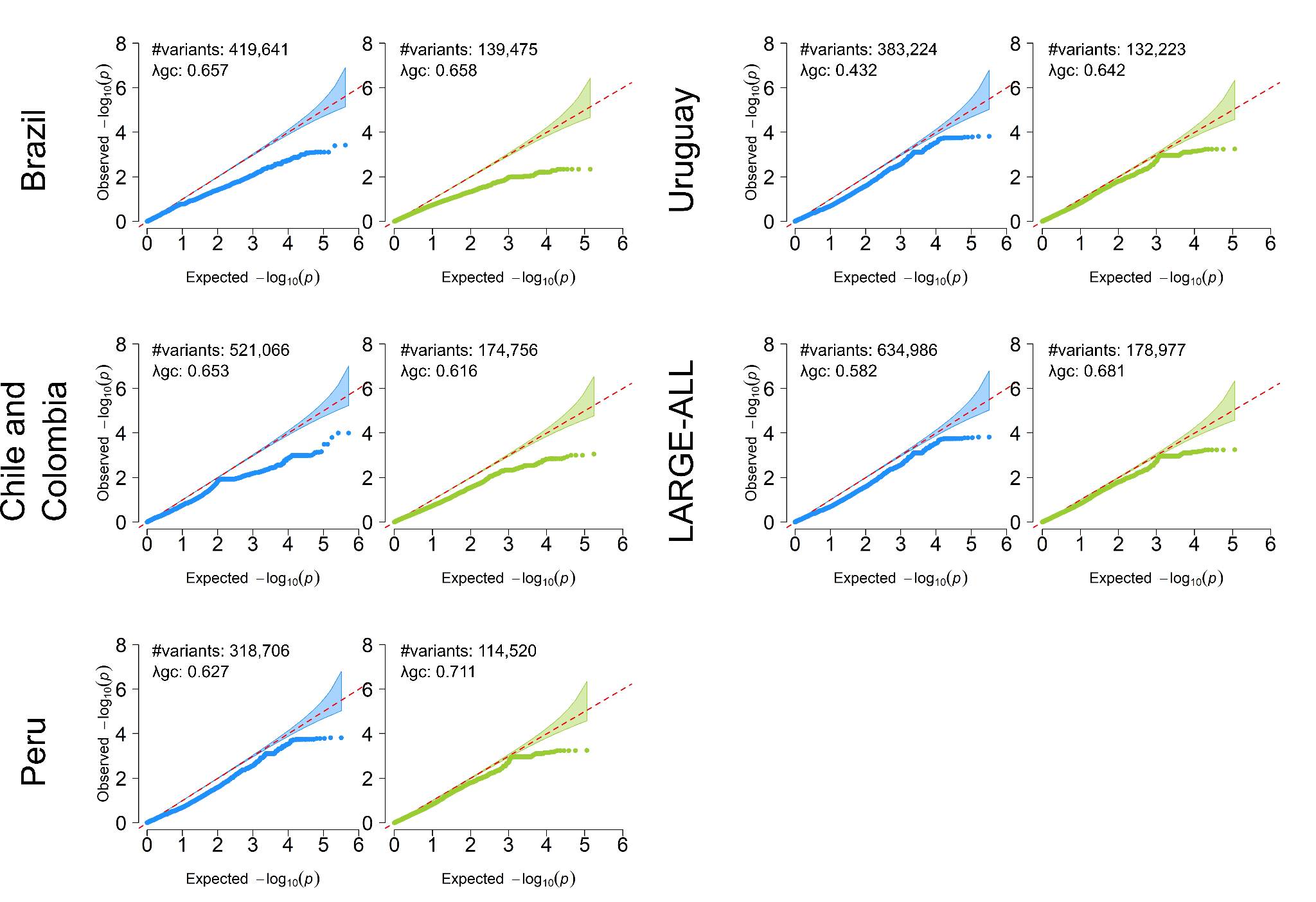

##### Figure S9: QQPlots to all variants (in blue) and for homozygous variants only (in green). The difference between the number of variants exists because some variants, after removing the heterozygous, became monomorphic. λgc is the genomic inflation factor.

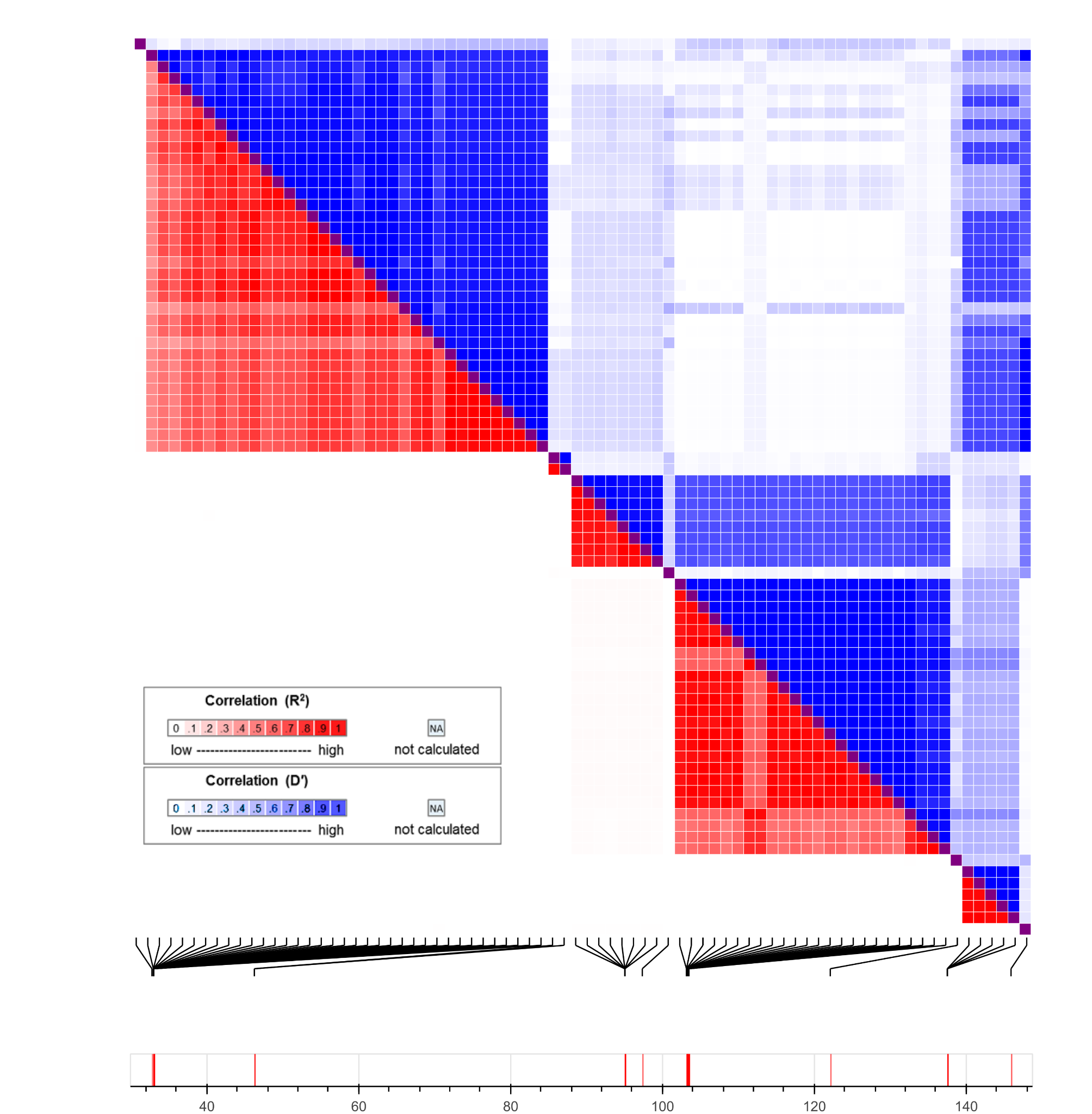

##### Figure S10: LD matrix for all the candidate variants. The colors represent the R² correlation (red) and coefficient of linkage disequilibrium (D’), in blue. We generated the plot using the LDmatrix Tool for our candidate variants, with the Admixed American Populations (MXL, PUR, CLM and PEL) from the 1000 Genomes Project as reference.

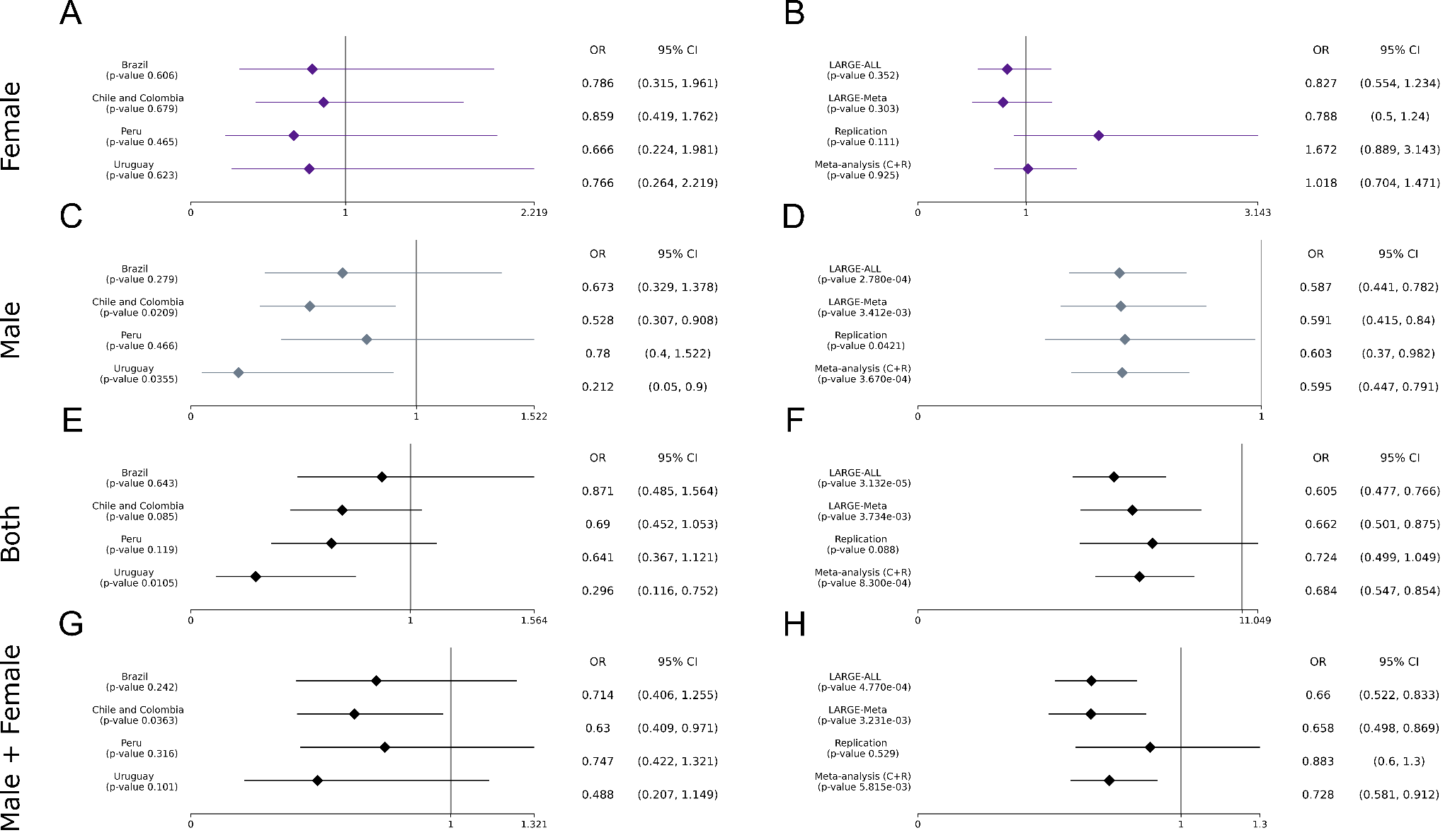

##### Figure S11: The Forest plot for the variant rs525496, the best variant in the replicated interval chr X: 103929276 - 104031236, for females (A and B), Males (C and D), Both (E and F) and Male + Female (G and H). On the left side we present the p-value, OR and 95% CI per cohort from discovery phase and on the right side we show the results from discovery phase (LARGE-Meta and LARGE-ALL), the replication and the meta-analysis between discovery phase and replication.

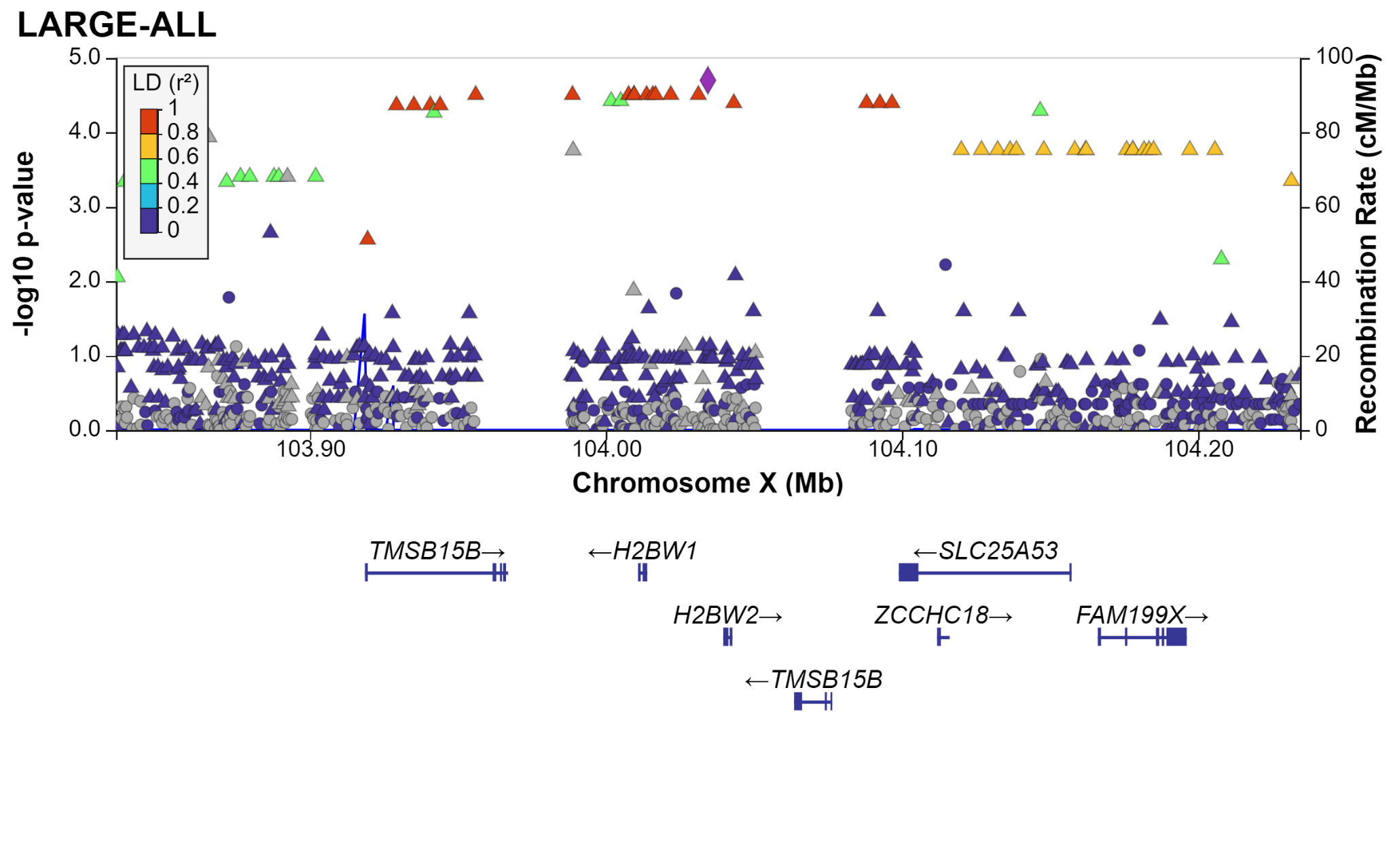

##### Figure S12: LocusZoom for the LARGE-ALL dataset for the replicated region between 103929276 - 104031236. The LD was calculated using the AMR populations.

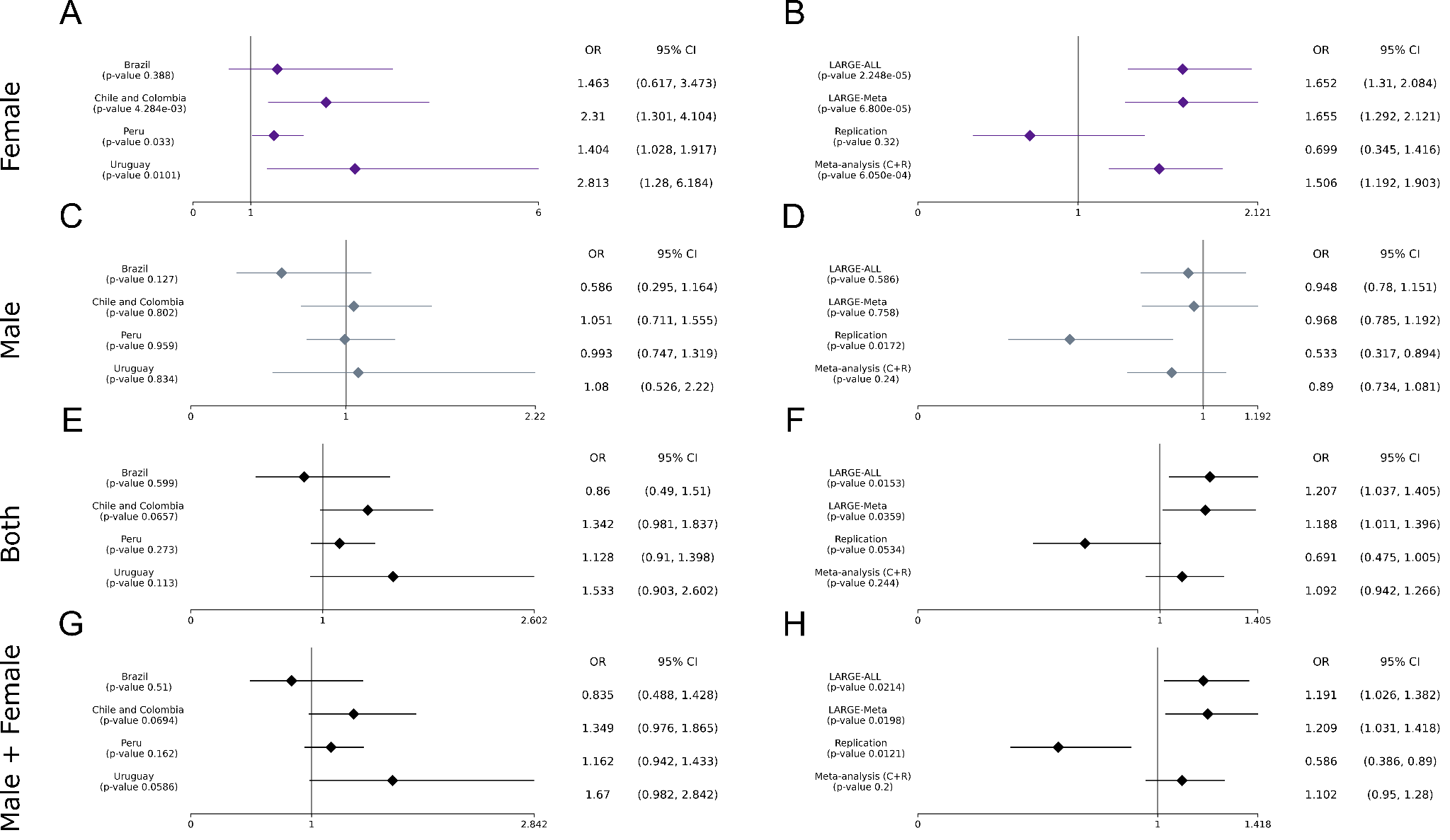

##### Figure S13: The Forest plots for variant rs112103361, the best inside the replicated interval chrX:95074293 - 95823918, for females (A and B), Males (C and D), Both (E and F) and Male + Female (G and H). On the left side we present the p-value, OR and 95% CI per cohort from discovery phase and on the right side we show the results from discovery phase (LARGE-Meta and LARGE-ALL), the validation and the meta-analysis between discovery phase and validation.

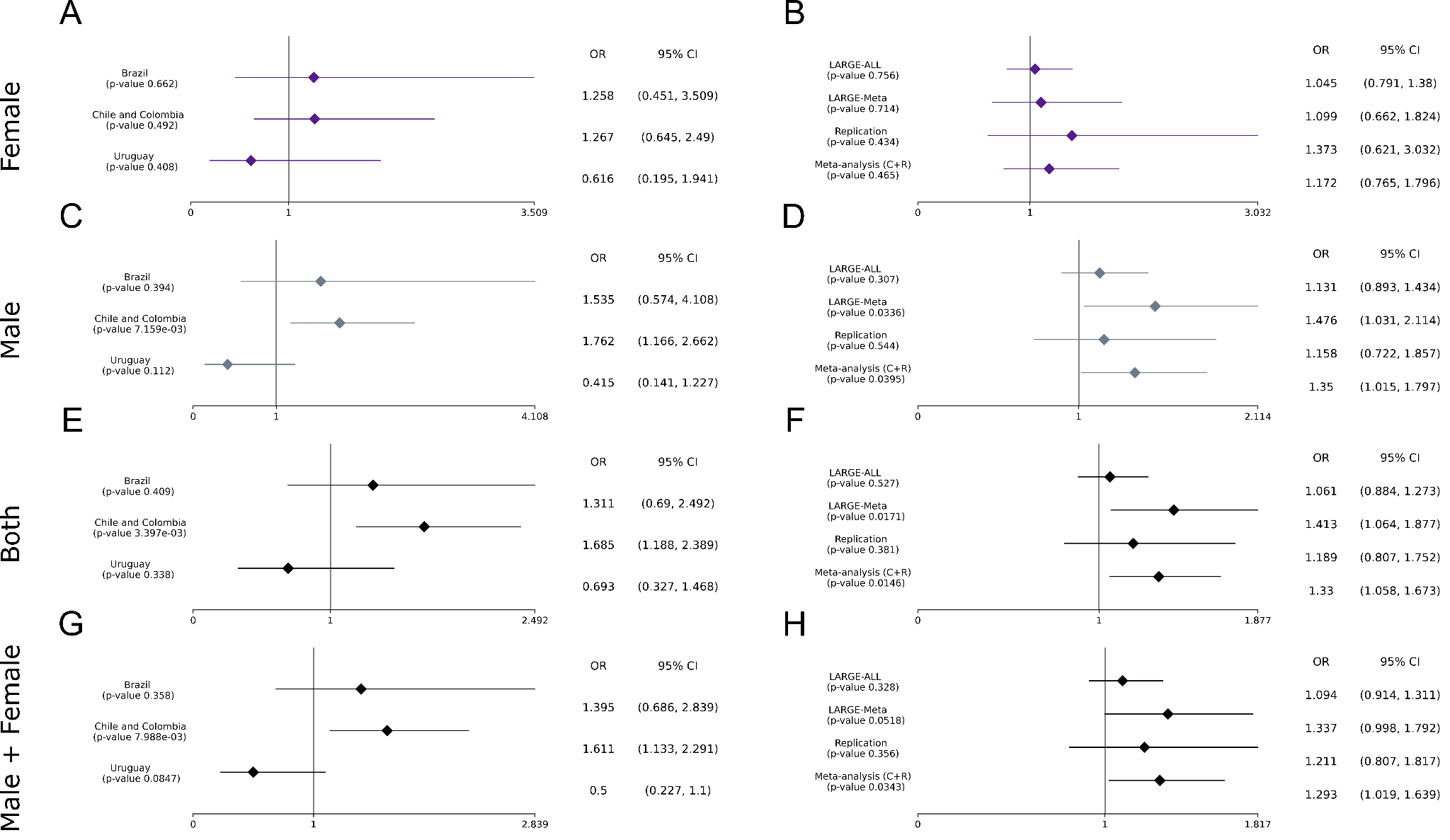

##### Figure S14: The Forest plot for variant rs28602900 for females (A and B), Males (C and D), Both (E and F) and Male + Female (G and H). On the left side we present the p-value, OR and 95% CI per cohort from discovery phase and on the right side we show the results from discovery phase (LARGE-Meta and LARGE-ALL), the replication and the meta-analysis between discovery phase and validation.

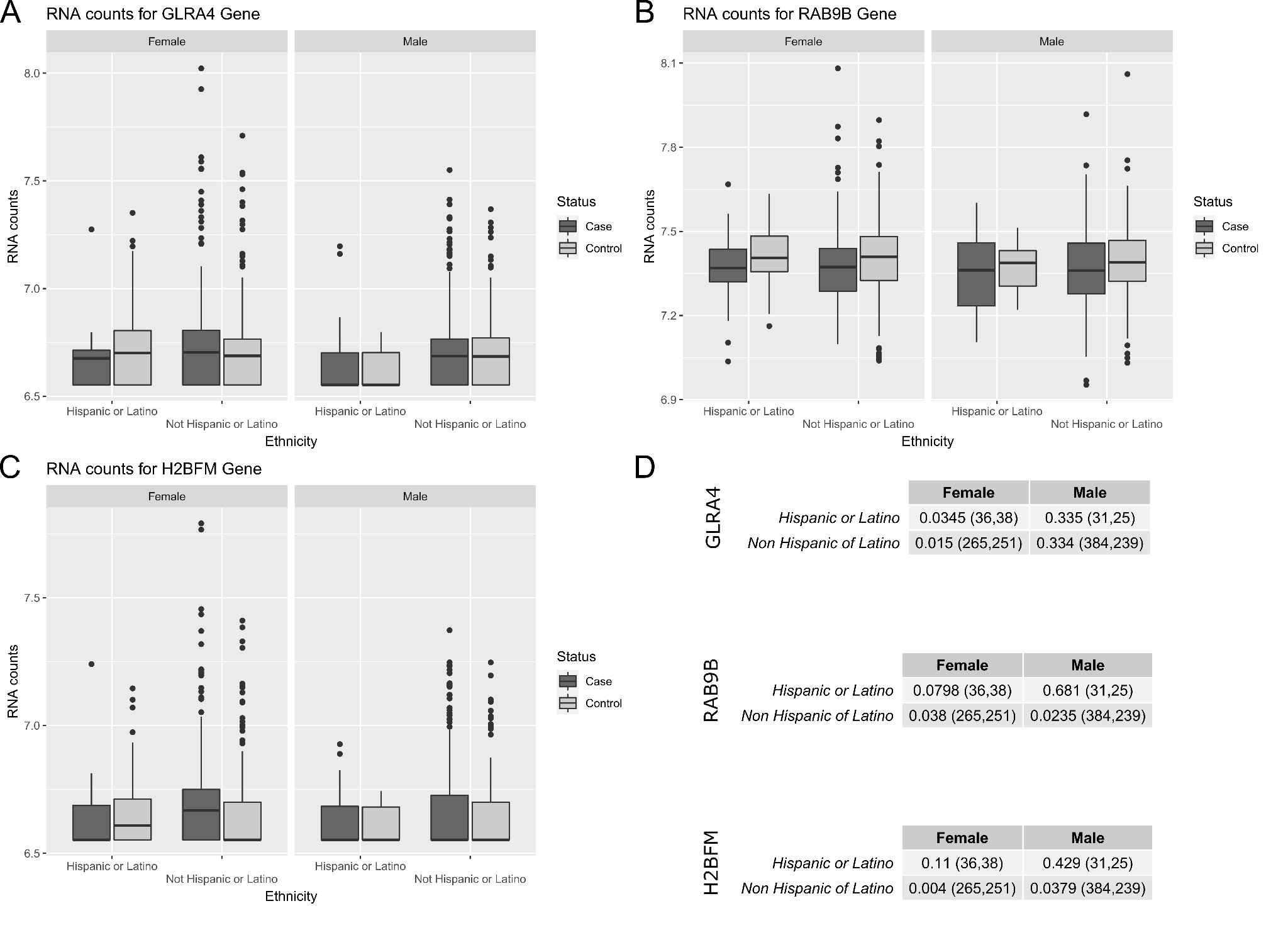

##### **Figure S15:** Distribution for RNA counts from PPMI project for the eQTL genes (A) GLRA4, (B) RAB9B, (C) HBFM segregated by sex and ethnicity. In (D) we show the p-value for a t-test between RNA counts for cases and controls. The number of cases and controls are shown in parenthesis.

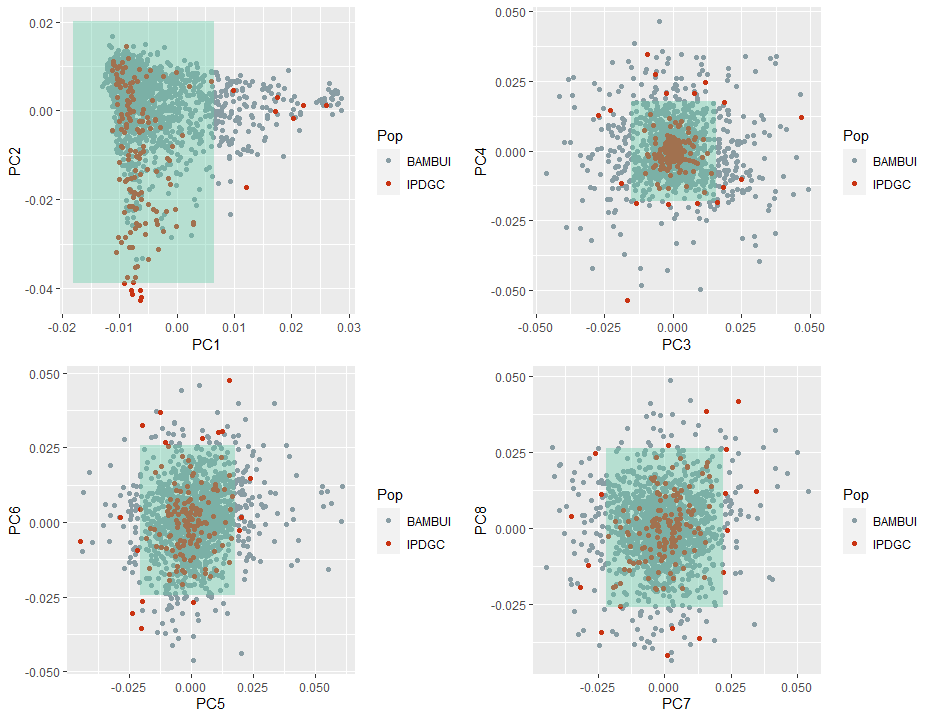

##### Figure S16: The IPDC PCA to select samples from the Bambuí dataset.

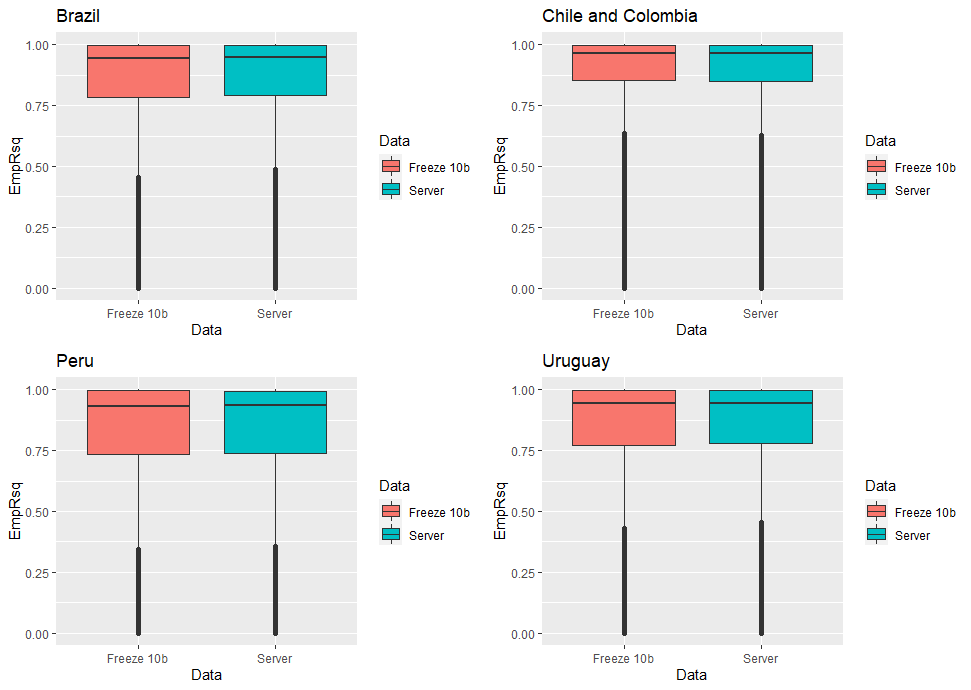

##### Figure S17: The EmpR and EmpRsq per cohort using TOPMed Freeze10b and the TOPMed Imputation Server

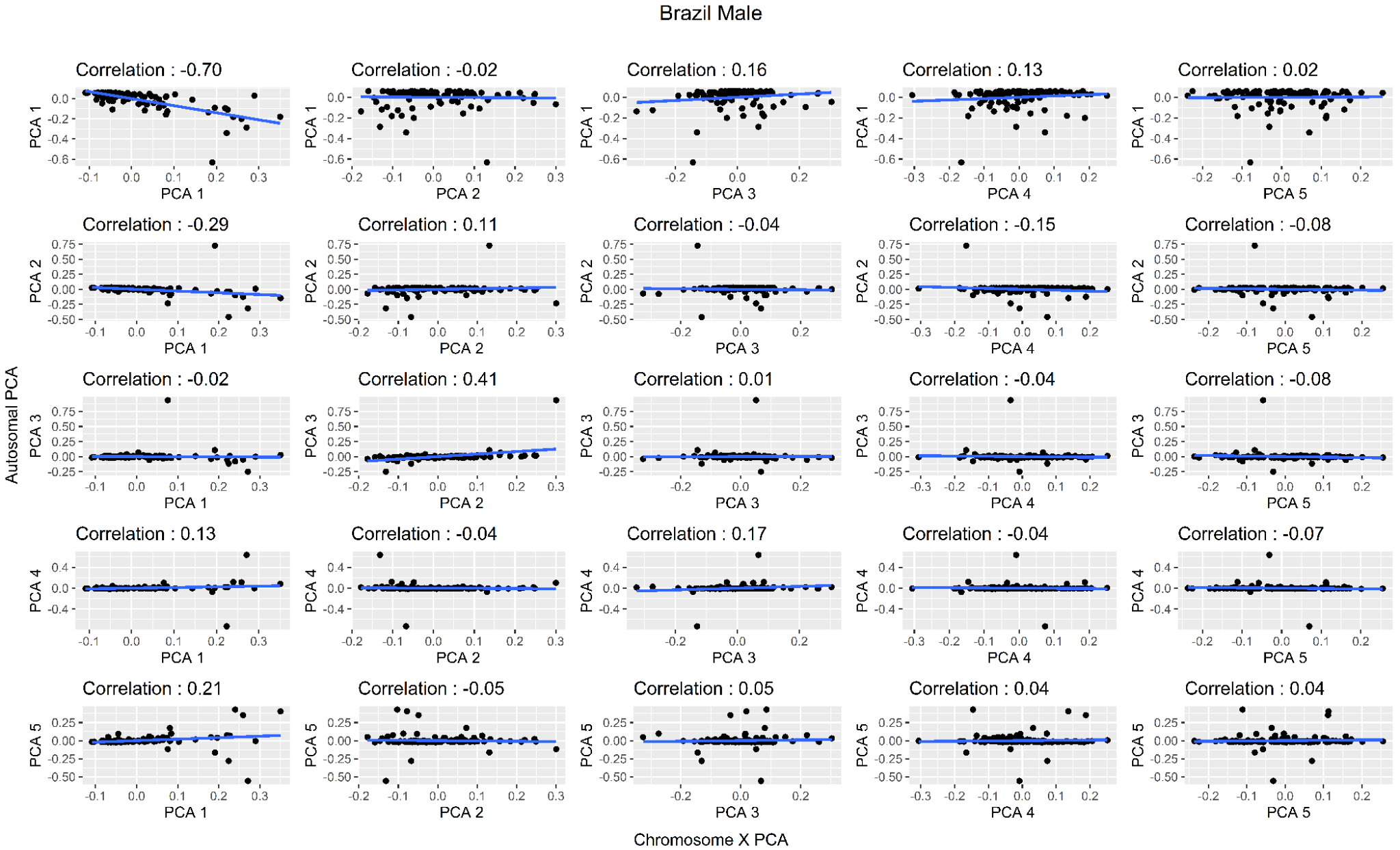

##### Figure S18 : Correlation between autosomal PCA (Y-axis) and the X-chromosome PCA (X-axis) for the Brazil Male dataset

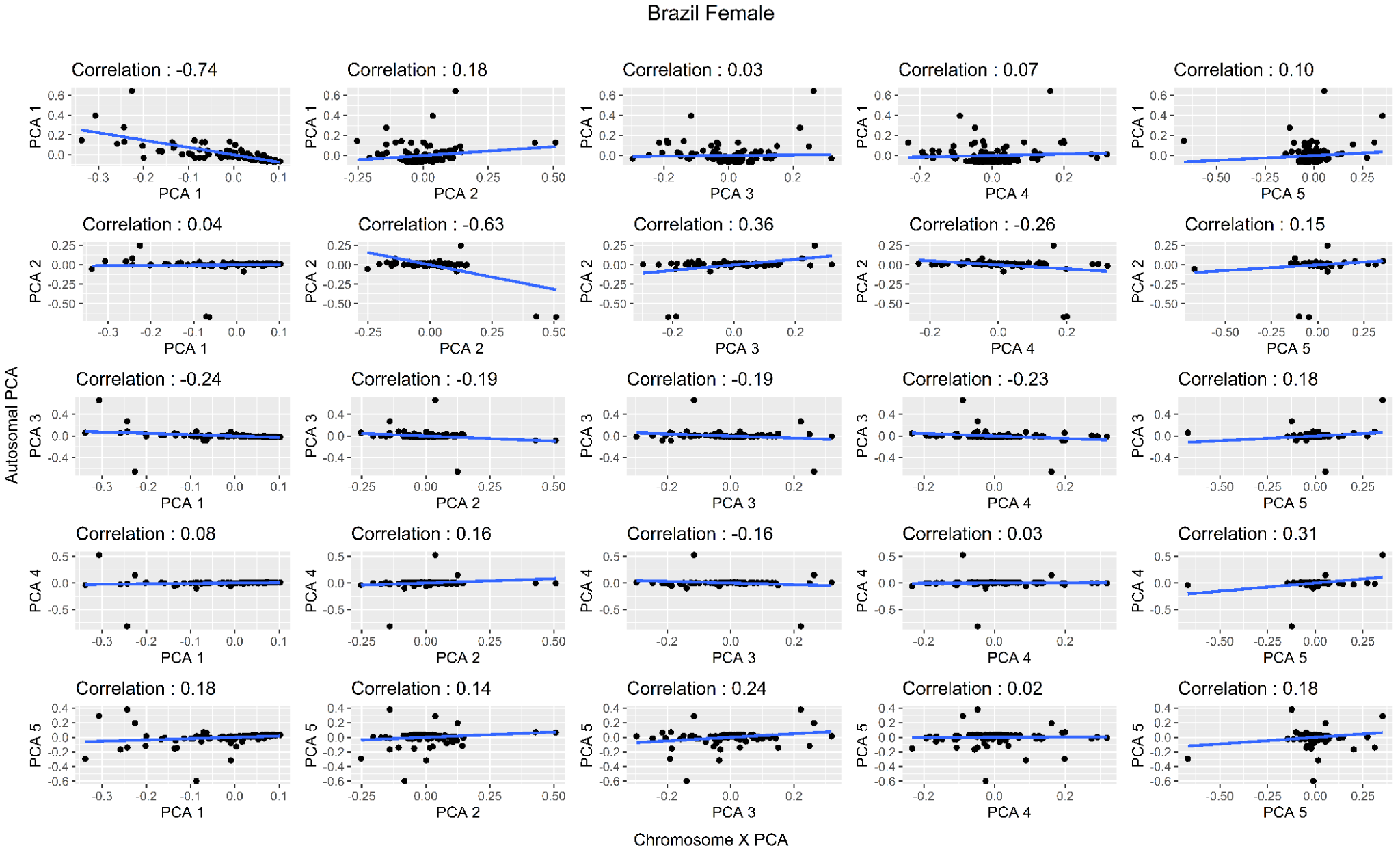

##### Figure S19 : Correlation between autosomal PCA (Y-axis) and the X-chromosome PCA (X-axis) for the Brazil Female dataset

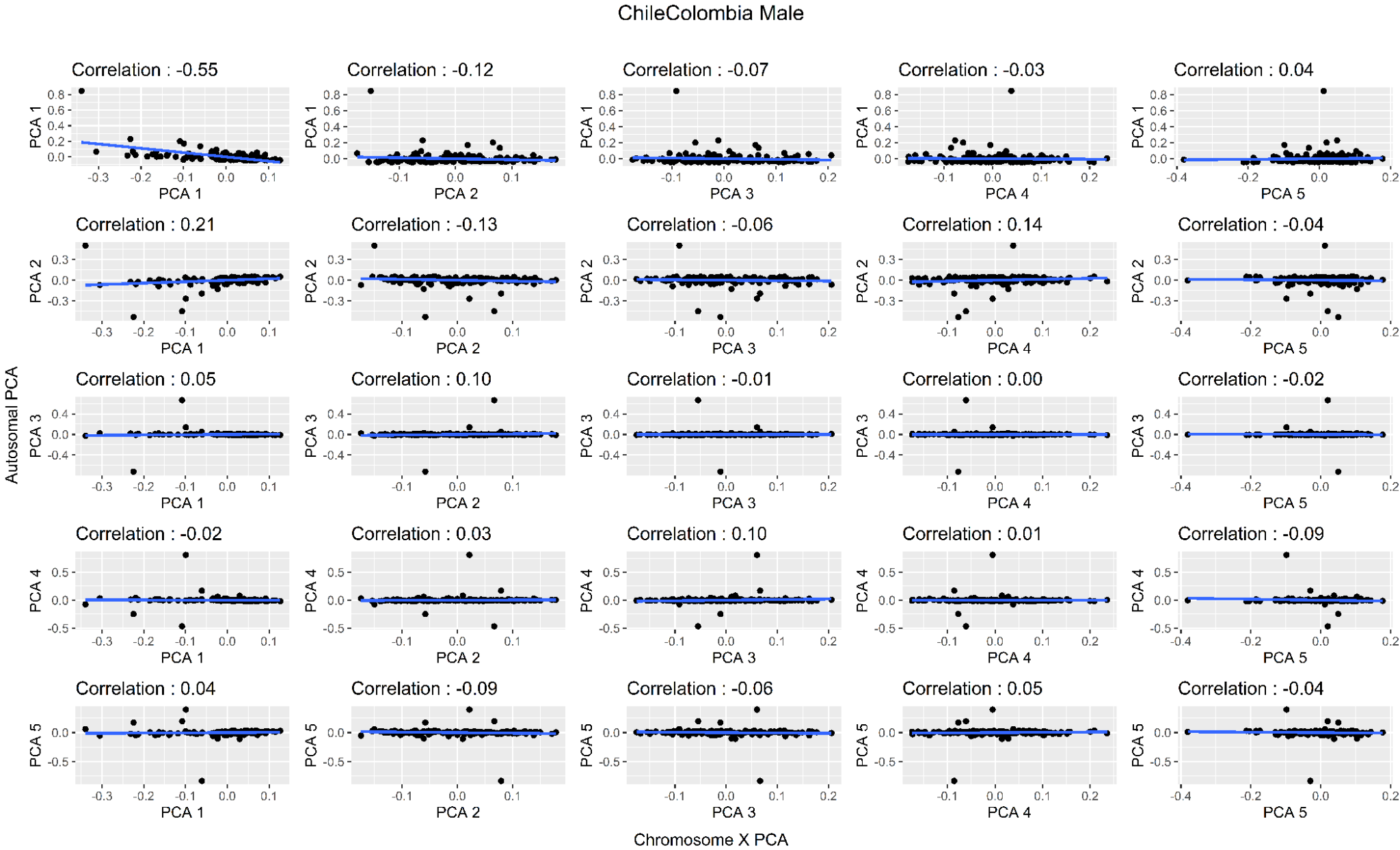

##### Figure S20 : Correlation between autosomal PCA (Y-axis) and the X-chromosome PCA (X-axis) for the Chile and Colombia Male dataset

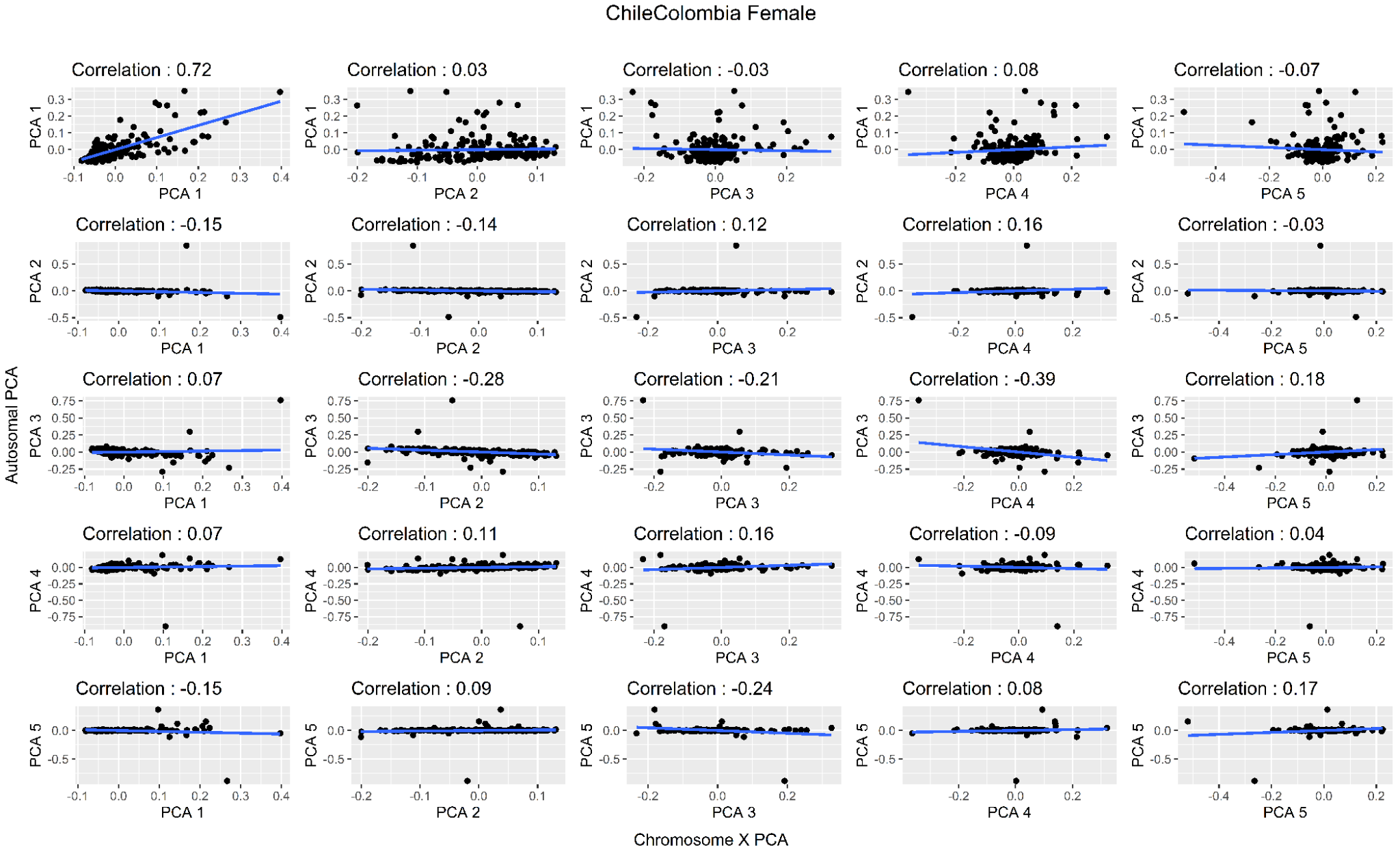

##### Figure S21 :Correlation between autosomal PCA (Y-axis) and the X-chromosome PCA (X-axis) for the Chile and Colombia Female dataset

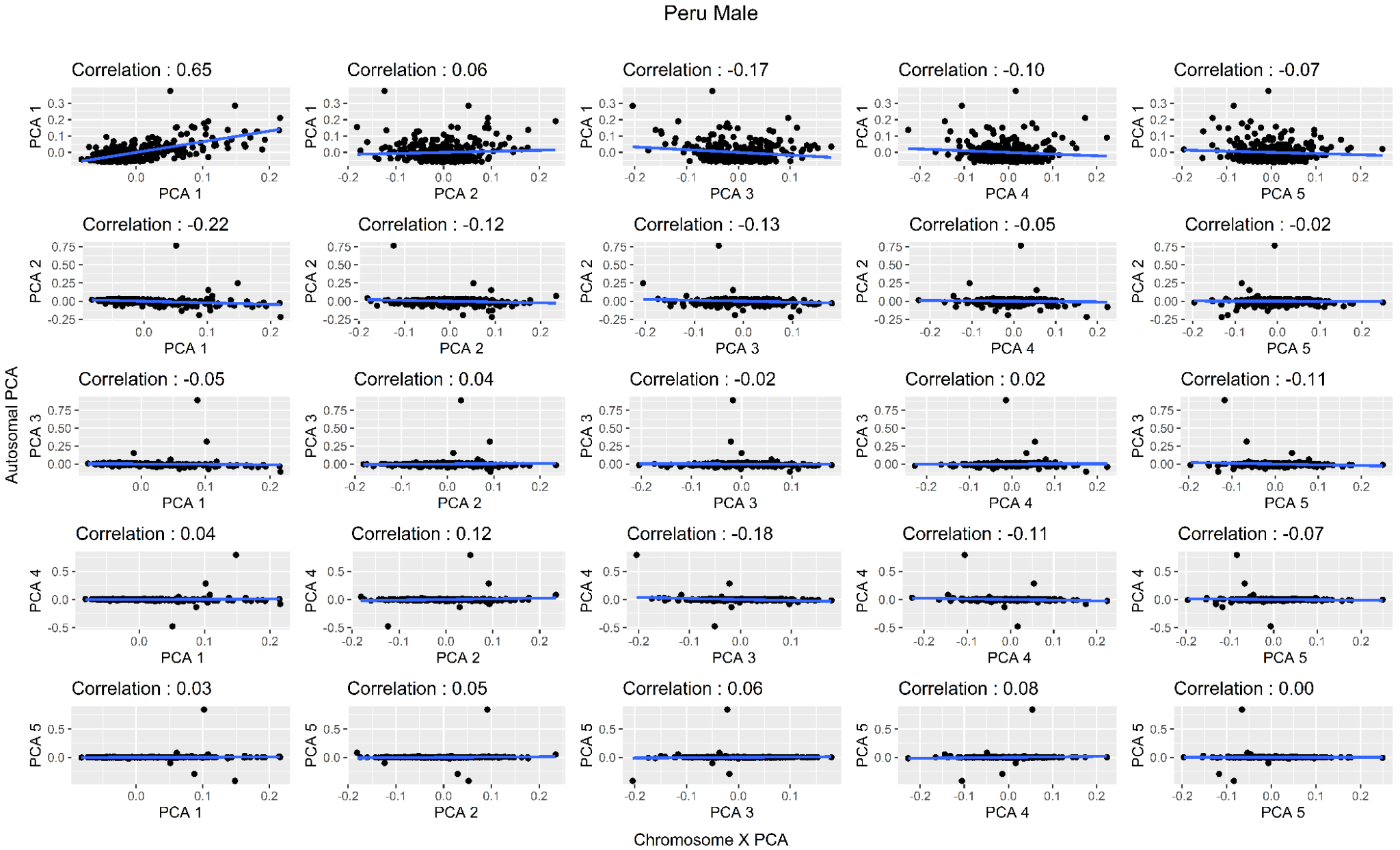

##### Figure S22: Correlation between autosomal PCA (Y-axis) and the X-chromosome PCA (X-axis) for the Peru Male dataset

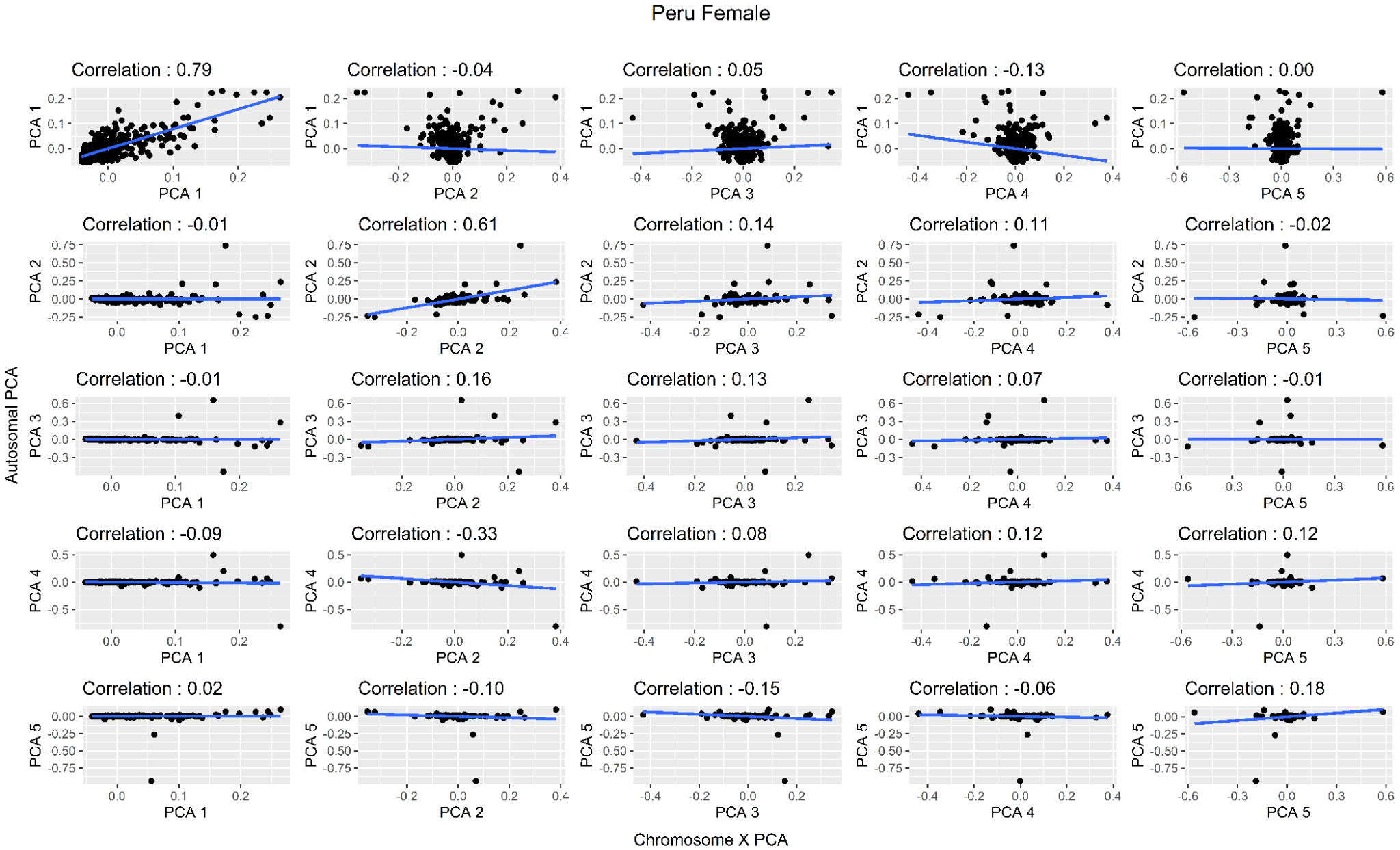

##### Figure S23 : Correlation between autosomal PCA (Y-axis) and the X-chromosome PCA (X-axis) for the Peru Female dataset

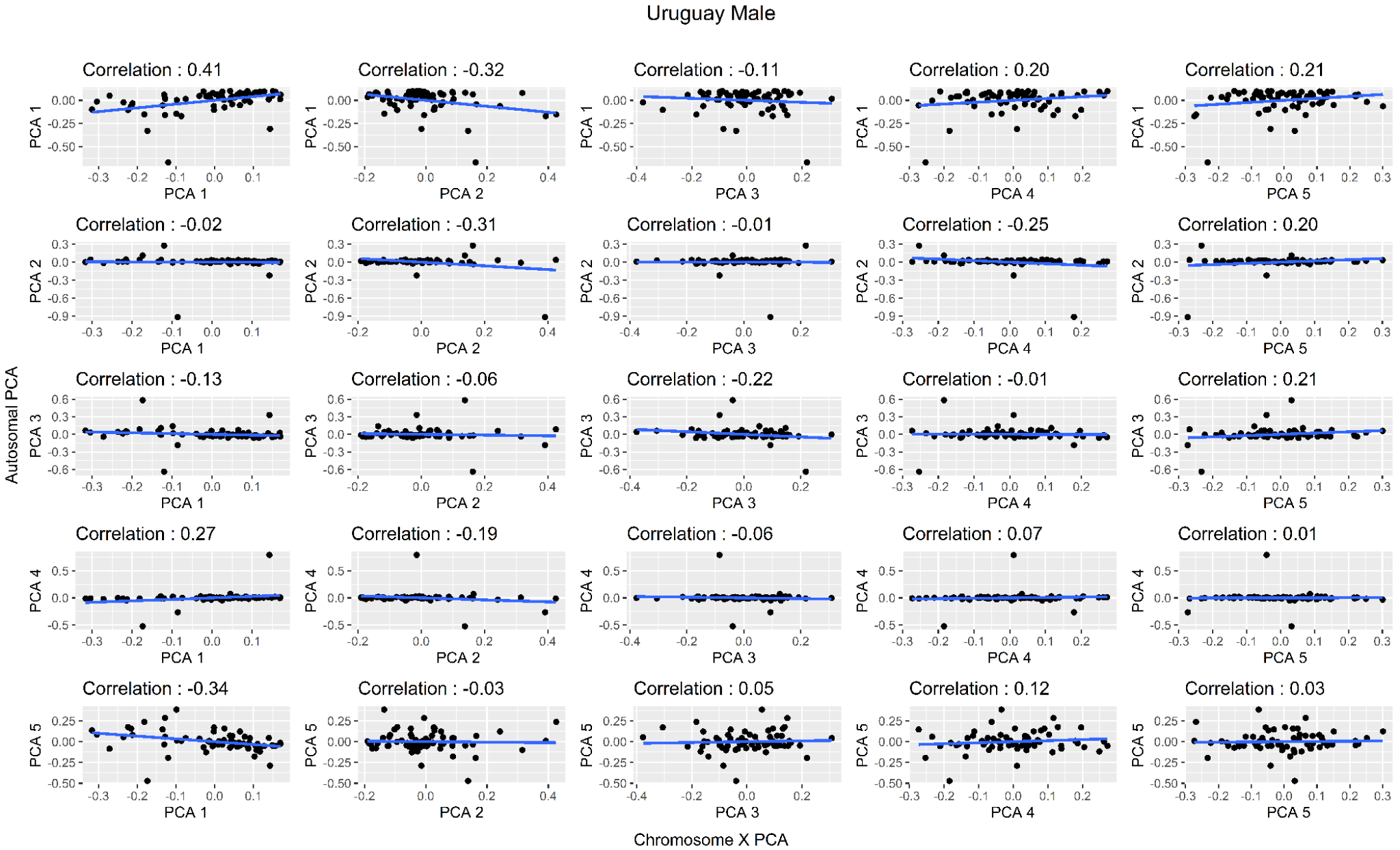

##### Figure S24 : Correlation between autosomal PCA (Y-axis) and the X-chromosome PCA (X-axis) for the Uruguay Male dataset

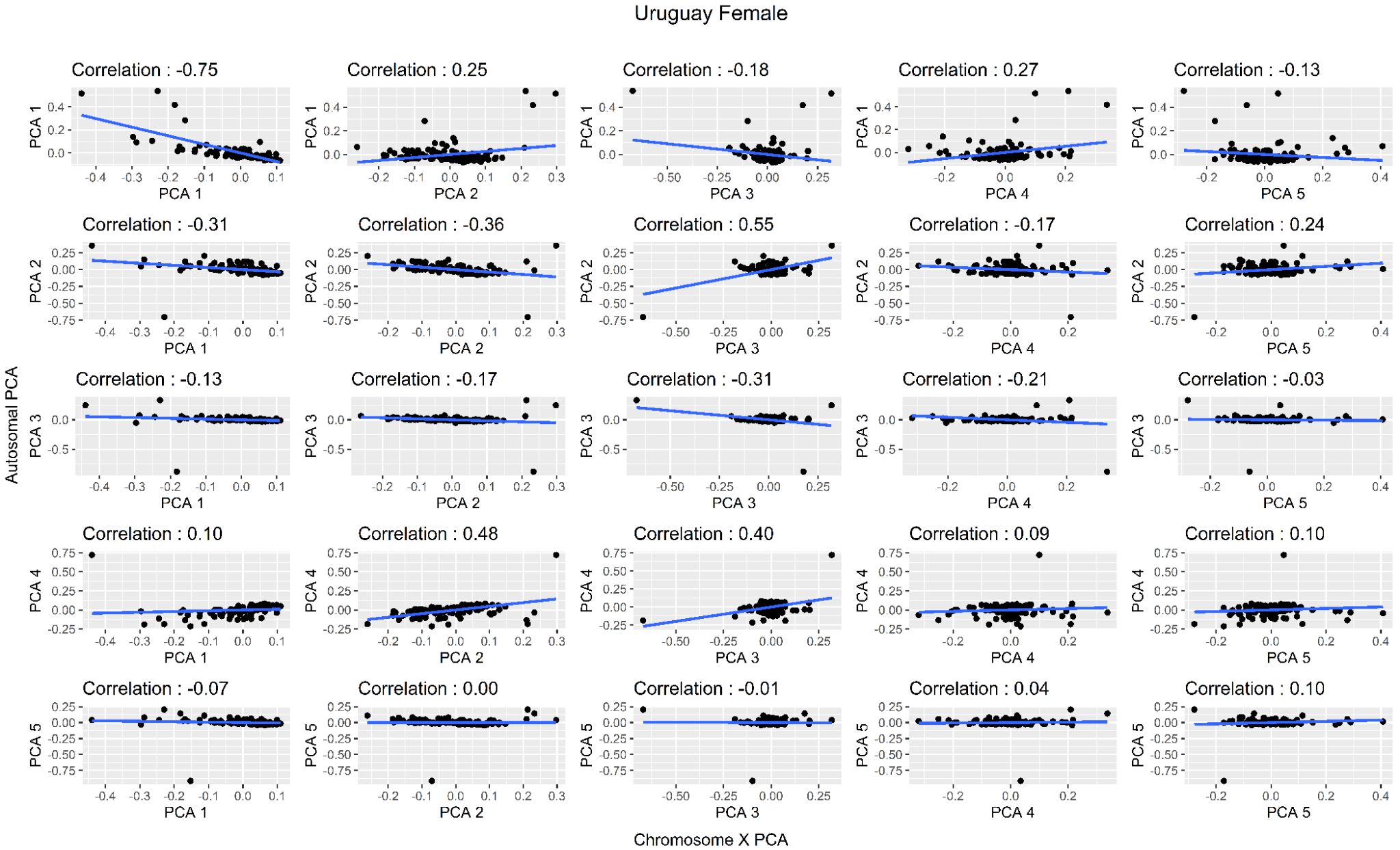

##### Figure S25 : Correlation between autosomal PCA (Y-axis) and the X-chromosome PCA (X-axis) for the Uruguay Female dataset

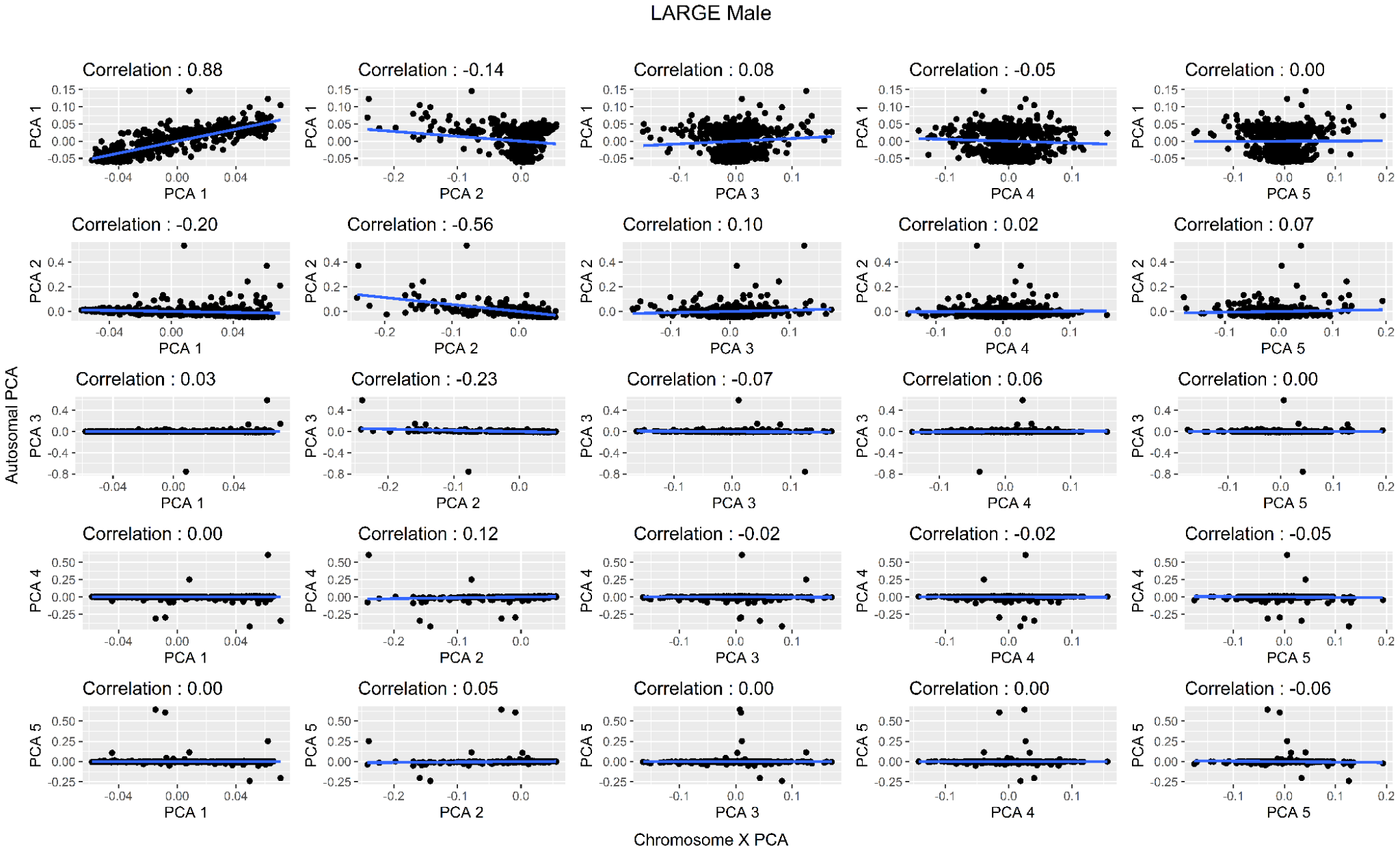

##### Figure S26 : Correlation between autosomal PCA (Y-axis) and the X-chromosome PCA (X-axis) for the LARGE-PD Male dataset

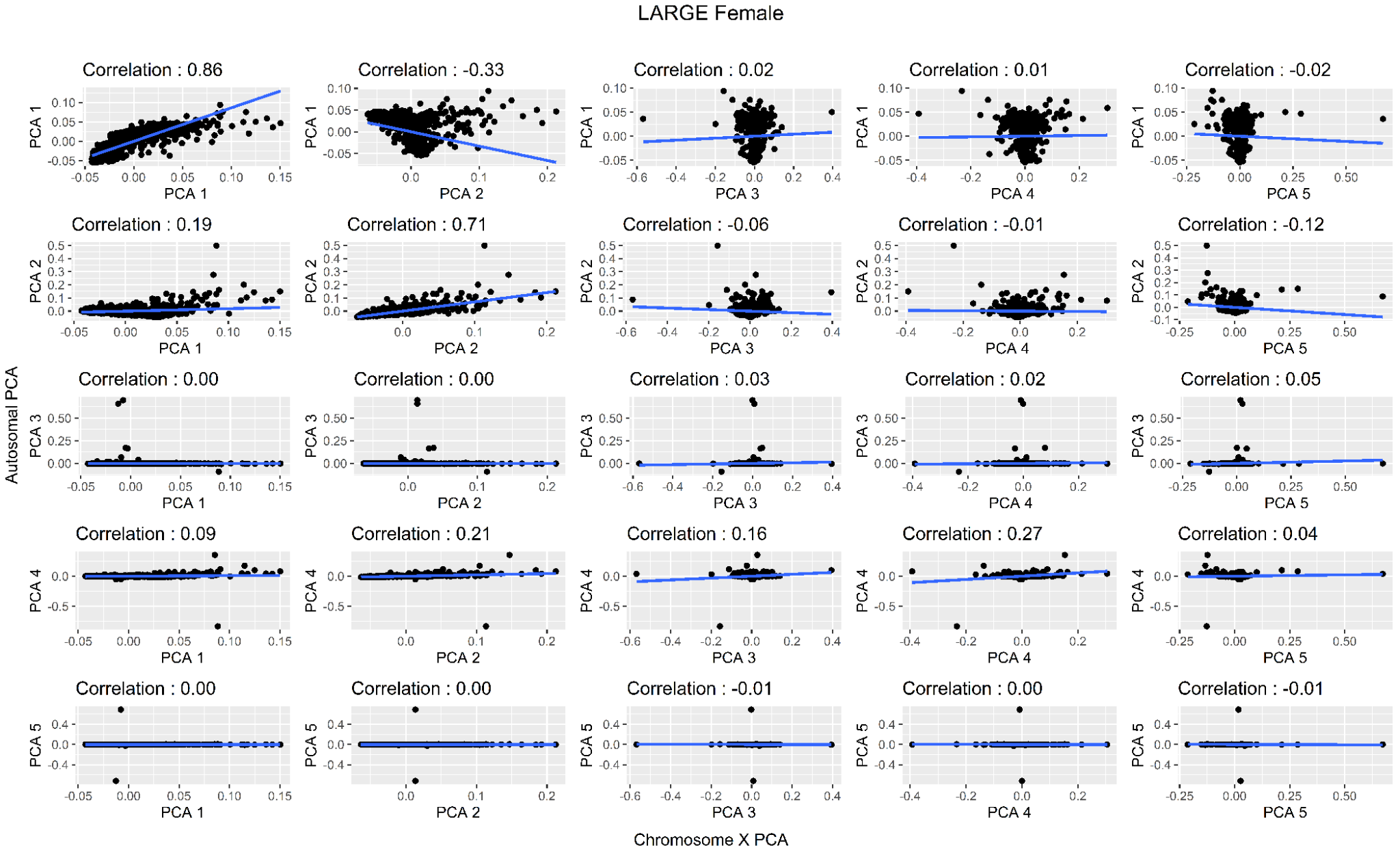

##### Figure S27:Correlation between autosomal PCA (Y-axis) and the X-chromosome PCA (X-axis) for the LARGE-PD Female dataset
